# Large tandem duplications in cancer result from transcription and DNA replication collisions

**DOI:** 10.1101/2023.05.17.23290140

**Authors:** Yang Yang, Michelle L. Badura, Patrick C. O’Leary, Henry M. Delavan, Troy M. Robinson, Emily A. Egusa, Xiaoming Zhong, Jason T. Swinderman, Haolong Li, Meng Zhang, Minkyu Kim, Alan Ashworth, Felix Y. Feng, Jonathan Chou, Lixing Yang

## Abstract

Despite the abundance of somatic structural variations (SVs) in cancer, the underlying molecular mechanisms of their formation remain unclear. Here, we use 6,193 whole-genome sequenced tumors to study the contributions of transcription and DNA replication collisions to genome instability. After deconvoluting robust SV signatures in three independent pan-cancer cohorts, we detect transcription-dependent replicated-strand bias, the expected footprint of transcription-replication collision (TRC), in large tandem duplications (TDs). Large TDs are abundant in female-enriched, upper gastrointestinal tract and prostate cancers. They are associated with poor patient survival and mutations in *TP53*, *CDK12*, and *SPOP*. Upon inactivating CDK12, cells display significantly more TRCs, R-loops, and large TDs. Inhibition of G2/M checkpoint proteins, such as WEE1, CHK1, and ATR, selectively inhibits the growth of cells deficient in CDK12. Our data suggest that large TDs in cancer form due to TRCs, and their presence can be used as a biomarker for prognosis and treatment.

## Introduction

Genome instability is a hallmark of cancer. Somatic structural variations (SVs) are abundant in human cancers, especially in solid tumors ^1,2^. However, our knowledge of their underlying causes is rather limited. In breast, ovarian, prostate, and pancreatic cancers, small deletions and small tandem duplications (TDs) can be attributed to homologous recombination deficiency (HRD) caused by *BRCA1* or *BRCA2* mutations ^3,4^. In lymphoid cells, translocations can result from aberrant V(D)J recombination ^5,6^. Many other tumor types also have numerous somatic SVs ^2^, but their causes remain unclear. The molecular mechanisms underlying genome instability are of clinical significance. For example, individuals carrying germline *BRCA* mutations are at higher risk of breast and ovarian cancers, so it is recommended that cancer screening begins between the ages of 25-35 ^7^. More importantly, the genes and pathways associated with the formation of somatic SVs may be druggable. For example, cancers with HRD can be effectively treated by PARP inhibitors ^8,9^.

DNA replication stress is a major source of genome instability ^10,11^. The various types of stress include deregulated origin firing, a shortage of nucleotides/proteins, and stalled replication forks. Many studies have demonstrated that the activation of oncogenes, such as Ras, Myc, Mos, and Cyclin E, can promote replication stress, induce collapsed replication forks, and lead to DNA double strand breaks (DSBs) and SVs ^12–17^. Here, we focus on the collision between transcription and DNA replication machineries, which is unavoidable because both processes use the same DNA template (**Fig. 1a**). Unresolved head-on collisions result in an accumulation of R-loops, which form DNA breaks ^18^. If not repaired properly, the collision may lead to replication fork collapse and genome instability ^19^. Some very large genes associated with common fragile sites are hotspots for deletions due to transcription-replication collision (TRC) ^20^. Deletions, insertions, and point mutations can frequently form when TRCs are induced in a bacterial system ^21^. Furthermore, a recent study using population sequencing data revealed transcription- and replication-dependent mutational processes in the human germline ^22^. However, the roles of TRCs in genome instability in human cancers remain largely unknown. Moreover, identifying tumors with high levels of TRCs may be clinically actionable, since drugs targeting transcription ^23,24^, replication ^25^, and DNA damage repair ^26^ are available.

**Figure 1.**
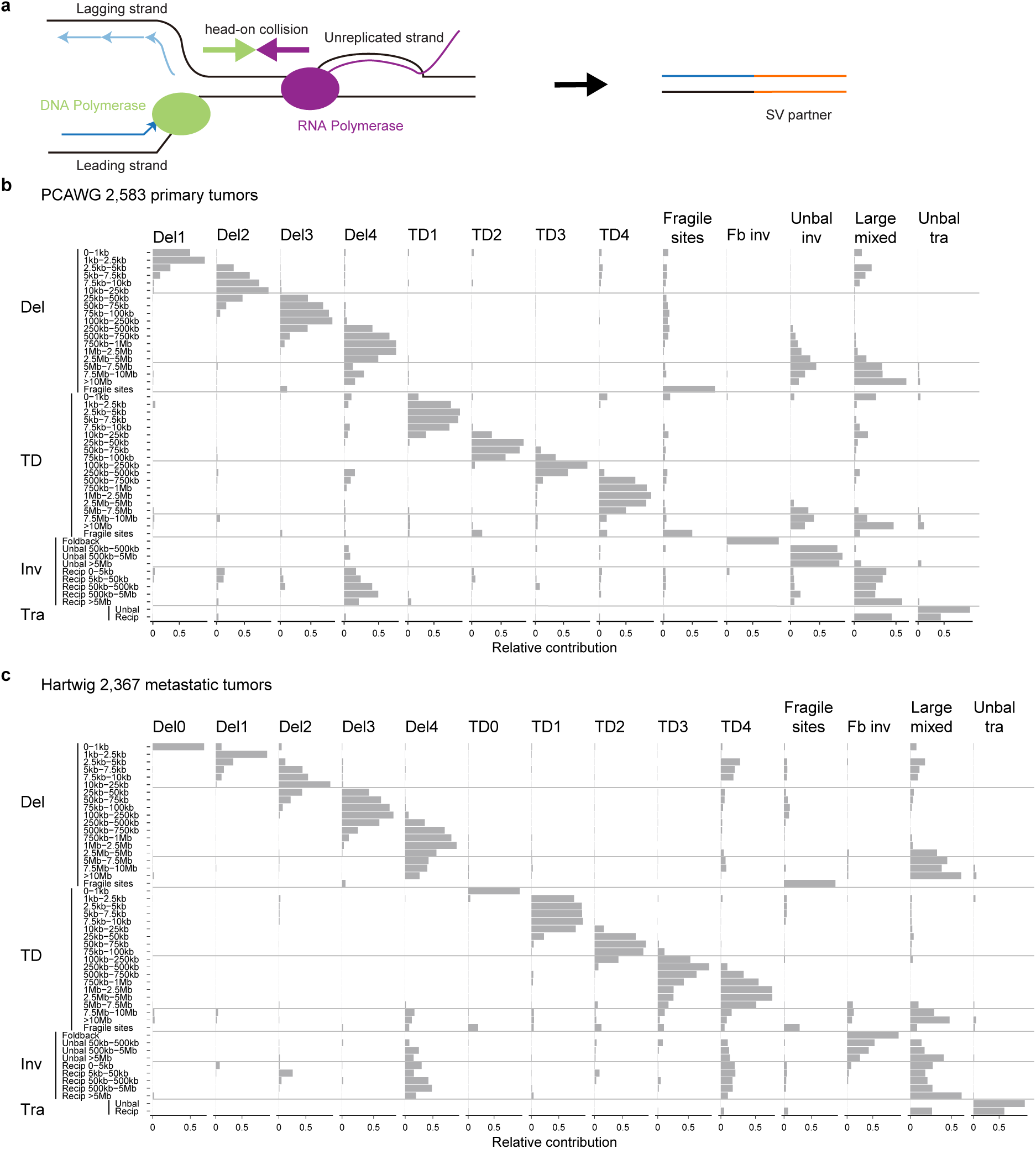
Transcription-replication collision and simple SV signatures. **a**, Diagram of the transcription and DNA replication collision producing SVs. The dark and light blue arrows in the left panel represent leading and lagging strands, respectively. The right panel illustrates an SV resulting from the collision. One of the replicated strands after the collision is ligated with a partner DNA fragment. The resulting SV can be either intra-chromosomal or inter-chromosomal, depending on the source of the SV partner. **b** and **c**, Simple SV signatures of 2,583 primary tumors in the PCAWG cohort (**b**) and 2,367 metastatic tumors in the Hartwig cohort (**c**). Simple SVs are classified into four broad categories (Del, deletion; TD, tandem duplication; Inv, inversion; and Tra, translocation) and further into 49 categories based on size. The y-axis shows the four broad categories and 49 categories. The x-axis shows 13 and 14 signatures for the two cohorts, respectively. The names of signatures are listed at the top. The lengths of the grey bars indicate the relative contributions of different categories to the corresponding signatures. The lengths of the grey bars for each category add up to 1.

Here, we used a total of 6,193 whole-genome sequenced tumors to perform a detailed analysis of somatic SVs. We revealed that large TDs result from TRCs and tumor cells with elevated TRCs can be selectively killed by multiple inhibitors targeting the G2/M checkpoint.

## Results

### Simple SV signatures in human cancers

Non-negative-matrix-factorization-(NMF)-based mutational signature decomposition is very effective at extracting signatures and inferring the underlying molecular mechanisms for somatic single nucleotide variants (SNVs) ^27,28^, copy number variations (CNVs) ^29,30^, SVs ^2,4^ and chromosomal instability ^31^. Recently, we have discovered six complex SV signatures in the Pan-Cancer Analysis of Whole Genomes (PCAWG) cohort and identified one signature related to circular extrachromosomal DNA (ecDNA) displaying bias in TRC regions ^32^. In the current study, we focus on TRCs in simple SVs.

We hypothesized that a subset of somatic simple SVs in cancer result from improperly repaired TRCs. Since NMF-based signatures can robustly capture molecular mechanisms, we used three large-scale pan-cancer whole-genome sequencing (WGS) cohorts to reliably infer simple SV signatures: the PCAWG cohort (2,583 primary tumors across 39 tumor types) ^33^, the Hartwig Medical Foundation cohort (Hartwig cohort thereafter, 2,367 metastatic tumors across 36 tumor types) ^34^, and the Personalized OncoGenomics cohort (POG570, 570 metastatic tumors across 26 tumor types) ^35^ (**Supplementary Table S1**). With a combined set of 5,520 tumors, our study is the largest pan-cancer SV signature study in human cancer to date to the best of our knowledge. In these three cohorts, there were a total of 1,025,587 somatic SVs (288,417 in PCAWG, 668,492 in Hartwig, and 68,678 in POG570). Previously, 12 SV signatures were identified in the PCAWG study ^2^, of which some complex SVs (i.e., chromothripsis) were excluded while other complex SVs (e.g., chromoplexy, cycle of templated insertions, chain of templated insertions, etc.) were included. Therefore, the previous PCAWG SV signatures were a mixture of simple and complex SVs. We have shown that complex SVs form via distinct mechanisms ^32^. Hence, to better capture the simple SV (deletions, TDs, inversions, and translocations) signatures, all types of complex SVs were excluded in our current study. As a result, there were 133,718, 303,905, and 41,014 simple SVs in the PCAWG, Hartwig, and POG570 cohorts, respectively. To better resolve the effect of SV size, we divided both deletions and TDs into 18 different size categories (**Fig. 1b** and Methods). All simple SVs were classified into 49 categories based on SV type and size, and SigProfilerExtractor ^36^ was used to deconvolute SV signatures. A total of 13 signatures were detected in the PCAWG cohort (**Fig. 1b** and **Supplementary Fig. S1**). There were four signatures of deletions and another four of TDs, as well as fragile site SVs, foldback inversions, unbalanced inversions, large mixed SVs (including large deletions, large TDs, reciprocal inversions, and reciprocal translocations), and unbalanced translocations. The deletion and TD signatures were named as “Del1” to “Del4” and “TD1” to “TD4” (**Fig. 1b**). Our 13 SV signatures matched the previous 12 signatures ^2^ very well (**Supplementary Fig. S2**) suggesting the robustness of NMF-based approach. The main differences were in deletion and TD signatures (**Supplementary Fig. S2**). This was due to differences in SV size categories and whether replication timing was used in signature decomposition. Our SV signatures were based on 18 size categories for deletions and TDs, whereas the previous study ^2^ only separated them into four size categories (i.e., <50 kb, 50 kb - 500 kb, 500 kb - 5 MB, and >5 MB). Our TD2 and TD3 signatures were best separated at 100 kb (**Fig. 1b**). Therefore, our SV size categories are more biologically relevant. Furthermore, we aimed to model strand bias regarding replication orientations (see next section), and we therefore avoided implementing replication timing in SV signature extraction. Another study reported 3 tandem duplicator phenotypes ^37^. Three of our four TD signatures (TD1, TD3, and TD4) corresponded to Group 1, 2, and 3 tandem duplicator phenotypes. In the Hartwig cohort, we identified 14 signatures (**Fig. 1c** and **Supplementary Fig. S3**) which were very similar to the PCAWG signatures (**Fig. 1b**). There were two additional signatures of very small deletions and very small TDs, namely “Del0” and “TD0”, which were likely due to the differences in SV detection algorithms. In the Hartwig cohort, the unbalanced inversions were captured by both foldback inversion and large mixed SV signatures rather than as a standalone signature in the PCAWG cohort (**Fig. 1b** and **1c**). In the POG570 cohort, the signatures (**Supplementary Fig. S4**) were also consistent with the PCAWG and Hartwig cohorts. In all three cohorts, reciprocal inversions and reciprocal translocations were captured by one signature, which suggested that they may form through a common molecular mechanism. In contrast, unbalanced inversions and unbalanced translocations were captured by distinct signatures, which suggested that they were generated via distinct mechanisms. Interestingly, in all three cohorts, a small fraction of the reciprocal inversions was captured by the large deletion signature Del4 (**Fig. 1b**, **1c** and **Supplementary Fig. S4a**), indicating possible shared molecular mechanisms.

The samples of the PCAWG, Hartwig, and POG570 cohorts were collected, processed, and sequenced at different facilities in different countries. Somatic SVs were called by different teams using different algorithms. The simple SV signatures deconvoluted from the three cohorts were highly reproducible. Therefore, our robust SV signatures represented mutational processes of simple SVs in cancer well, did not include any complex SVs, and offered a better resolution in SV size than the previous study ^2^. In the remainder of this study, we mapped simple SVs in primary tumors from additional cohorts to the 13 PCAWG signatures and simple SVs from metastatic tumors to the 14 Hartwig signatures.

### SV strand bias in transcription-replication collision regions

Here, we aim to identify SVs generated by improper repair of TRCs. The formation of these SVs should depend on both transcription and DNA replication. We found that small deletions, fragile site SVs, and foldback inversions were enriched in late replicated regions, whereas TDs and unbalanced translocations were enriched in early replicated regions (**Supplementary Fig. S5**). In addition, some SVs were enriched in transcribed genes ^38^ (**Supplementary Fig. S6a** and **S6b**). However, which SVs result from TRCs remains unclear. The conflicts between transcription and DNA replication can be resolved properly in normal cells by various DNA damage response pathways, such as the Fanconi Anemia pathway ^19^, which allows DNA replication to progress without error. If the collisions are not properly repaired, the collapsed replication forks can eventually lead to SVs (**Fig. 1a**). Importantly, these SVs will bear unique genetic footprints. First, the resulting SV breakpoints should be located in genes. Furthermore, when the replication fork collapses after TRC, one side of the collapsed site has been replicated, whereas the other side remains unreplicated. Specifically, **Fig. 1a** shows an example of a head-on collision in which replication moves from the left to the right and transcription moves from the right to the left. Upon collision, the DNA fragment on the left of the collision site has already been replicated (labelled as leading and lagging strands), whereas the DNA fragment on the right side has not. Since the replicated strand has two copies whereas the unreplicated strand has only one copy, one of the replicated strands (either leading strand or lagging strand) would be ligated to another repair partner and becomes an SV (**Fig. 1a** right panel). Thus, the replicated strand should be enriched in the SV junctions (**Fig. 1a**). Such dosage imbalance will lead to strand bias ^39^ of the resulting SVs. The SV type depends on the choice of repair partner. If the partner is a DNA end from another chromosome, the SV would be an inter-chromosomal translocation. If the partner is from the same chromosome, the SV type depends on the partner’s location and orientation relative to the collision site.

For each SV breakpoint, two partner DNA fragments are linked together. Each partner has an orientation, typically annotated as “+” or “-” to indicate whether the upstream or the downstream portion of the DNA is linked to its partner. Deletions have “+-” orientations whereas TDs have “- +” orientations (**Supplementary Fig. S7**). To detect the aforementioned SV strand bias, we first defined conserved replication orientations for all protein-coding genes using Repli-seq data from 14 cell lines derived from solid tissues (**Fig. 2a** and **Supplementary Fig. S8**). Then, we randomly shuffled all observed SVs in the tumors and tested for strand bias in simple SVs in the PCAWG and Hartwig cohorts with at least one breakpoint falling in protein-coding genes. By comparing observed SVs against the randomly shuffled ones, we found that the breakpoints of TD3 and TD4 were consistently enriched in the replicated strand in both cohorts (**Fig. 2b** and **2c**). Large TDs (TD3 and TD4) were abundant in female-enriched cancers (uterus, ovarian, and breast cancers), upper gastrointestinal tract cancers (esophageal and stomach cancers), prostate, and urothelial cancers (**Fig. 2d** and **2e**). Large TDs were also abundant in metastatic head and neck cancers, which include parts of the upper digestive tract as well (**Fig. 2e**). In breast cancer, basal and HER2+ subtypes carried more large TDs than luminal A and B subtypes (**Supplementary Fig. S9**). Using replication orientations determined from the breast cancer cell line MCF7 and the liver cancer cell line HepG2, the strand bias could be observed in TD3 and TD4 in four independent breast cancer cohorts (PCAWG, Hartwig, POG570, and BRCA-EU) and liver cancer (**Fig. 3a** and **3b**). TD3 and TD4 also displayed strand bias in three independent ovarian cancer cohorts (PCAWG, Hartwig, and OV-AU) (**Fig. 3c and 3d**), metastatic prostate, and esophageal cancers (**Fig. 3d**).

**Figure 2.**
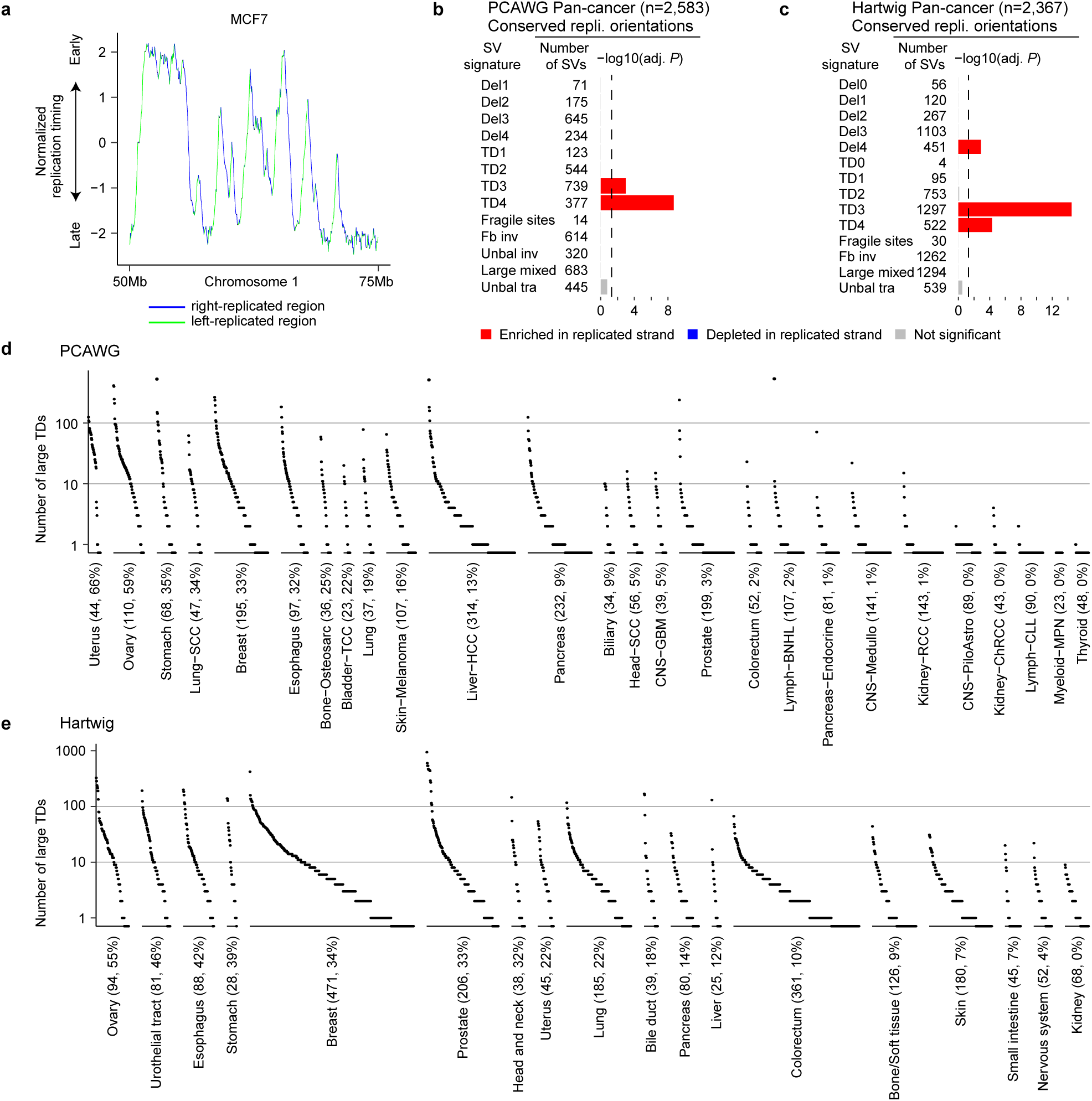
Pan-cancer replicated-strand bias and large TD frequencies across tumor types. **a**, An example of normalized replication timing of MCF7 cell line. The blue and green curves represent right- and left-replicated regions (sharp transition regions). **b** and **c**, Replicated-strand bias for primary tumors in the PCAWG cohort (**b**) and metastatic tumors in the Hartwig cohort (**c**). Each signature and its number of observed breakpoints are shown on the y-axis. *P* values are calculated by Fisher’s exact test. The x-axis shows Bonferroni-adjusted *P* values. The dashed black line on the x-axis represents a 0.05 adjusted *P* value cutoff. Conserved replication orientation is used for both cohorts. **d** and **e**, The numbers of large TDs in the PCAWG (**d**) and Hartwig (**e**) cohorts. Black dots represent individual tumors grouped by tumor type. Tumor types are annotated at the bottom of the plots. The two numbers after tumor type names are sample size and the proportion of samples with >= 10 large TDs in the corresponding tumor types. Tumor types with at least 20 tumors are shown and are sorted by the proportion of samples with >= 10 large TDs.

**Figure 3.**
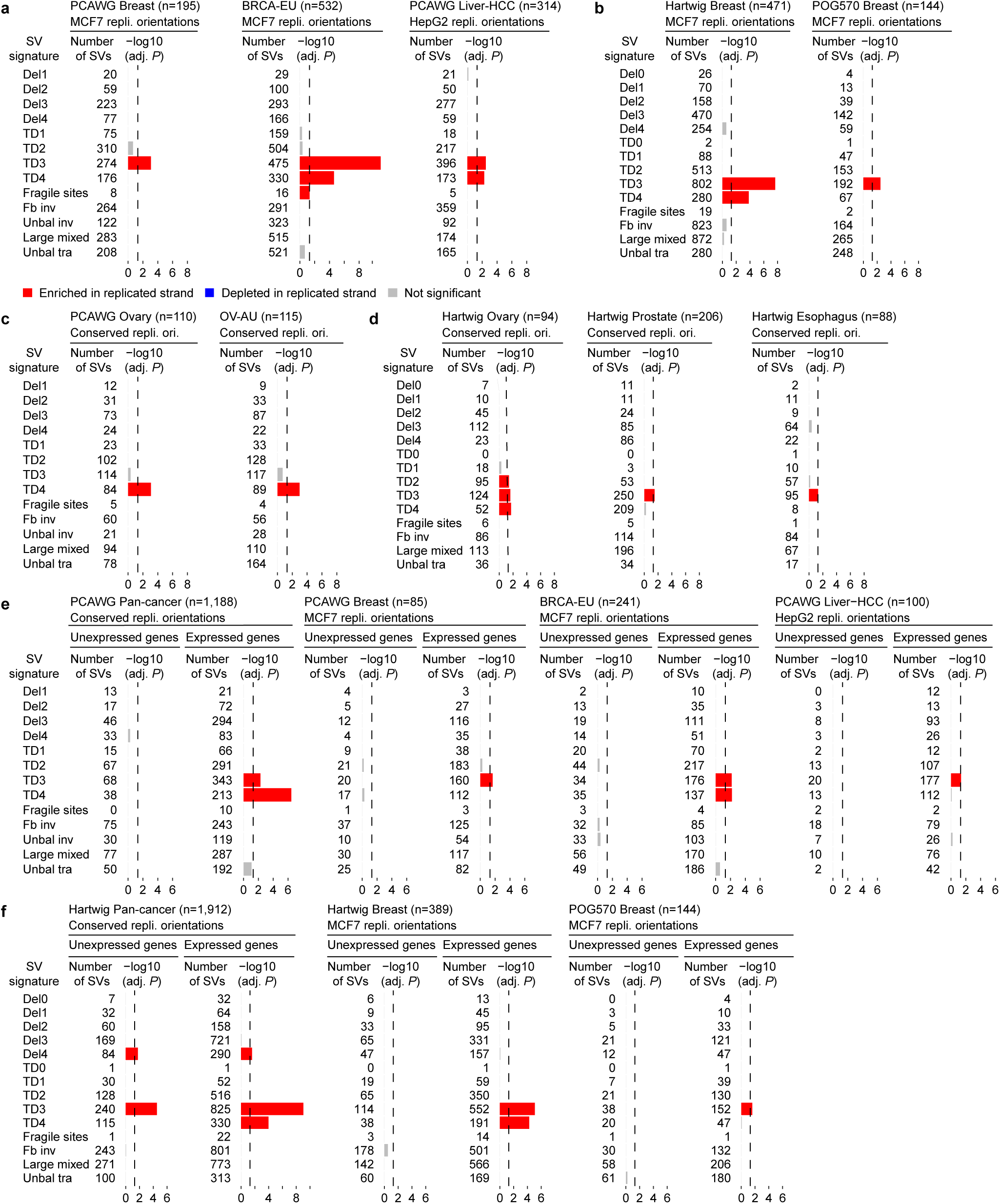
Tumor-type-specific and transcription-dependent replicated-strand bias. **a**, Replicated-strand bias in primary breast and liver cancers using replication orientations defined from the MCF7 and HepG2 cell lines. **b**, Replicated-strand bias in metastatic breast cancers using replication orientations defined from the MCF7 cell line. **c**, Replicated-strand bias in primary ovarian and prostate cancers. **d**, Replicated-strand bias in metastatic ovarian, prostate, and esophageal cancers. **e** and **f**, Replicated-strand bias in unexpressed genes and expressed genes for primary (**e**) and metastatic (**f**) tumors. For all panels, the tested tumor type, sample size and source of replication orientations are annotated at the top of each plot. *P* values are calculated by Fisher’s exact test. The x-axis shows Bonferroni-adjusted *P* values.

If large TDs result from TRCs, we expect the strand bias to be dependent on gene expression. In unexpressed genes, we did not observe strand bias in TD3 or TD4 in the PCAWG and Hartwig cohorts, whereas strand bias was clearly evident in expressed genes (**Fig. 3e** and **3f**). The same trend could be observed in four independent breast cancer cohorts (PCAWG, Hartwig, POG570, and BRCA-EU) and liver cancer using tissue-specific replication orientations (**Fig. 3e** and **3f**). The expression-dependent strand bias was also evident in two independent ovarian cancer cohorts (**Supplementary Fig. S10a**). It is possible that gene expression is the consequence of SVs rather than the cause. We therefore used gene expression from normal tissues and still observed the expression-dependent replicated-strand bias in TD3 and TD4 (**Supplementary Fig. S10b** and **S10c**). Taken together, we observed robust transcription-dependent replicated-strand bias in large TDs across multiple independent cancer cohorts.

### Replicated-strand bias as an independent effect in large TDs

Next, we sought to assess the effect of replication in strand bias of large TDs. There are two types of TRCs: head-on (transcription and replication in the opposite orientations, **Fig. 1a**) and co-directional (transcription and replication in the same orientations) collisions. If large TDs result from TRCs, we expect the replicated-strand bias to be observed in both types of collisions. Indeed, in the PCAWG and Hartwig cohorts, TD3 and TD4 displayed replicated-strand bias in both head-on and co-directional collisions (**Fig. 4a** and **4b**). The same was observed in BRCA-EU and Hartwig breast cancer cohorts when replication orientations were annotated from MCF7 cells (**Fig. 4a** and **4b**). These results suggest that the replicated-strand bias in large TDs depends on replication but not on the orientation (i.e., head-on vs co-directional) of collision.

**Figure 4.**
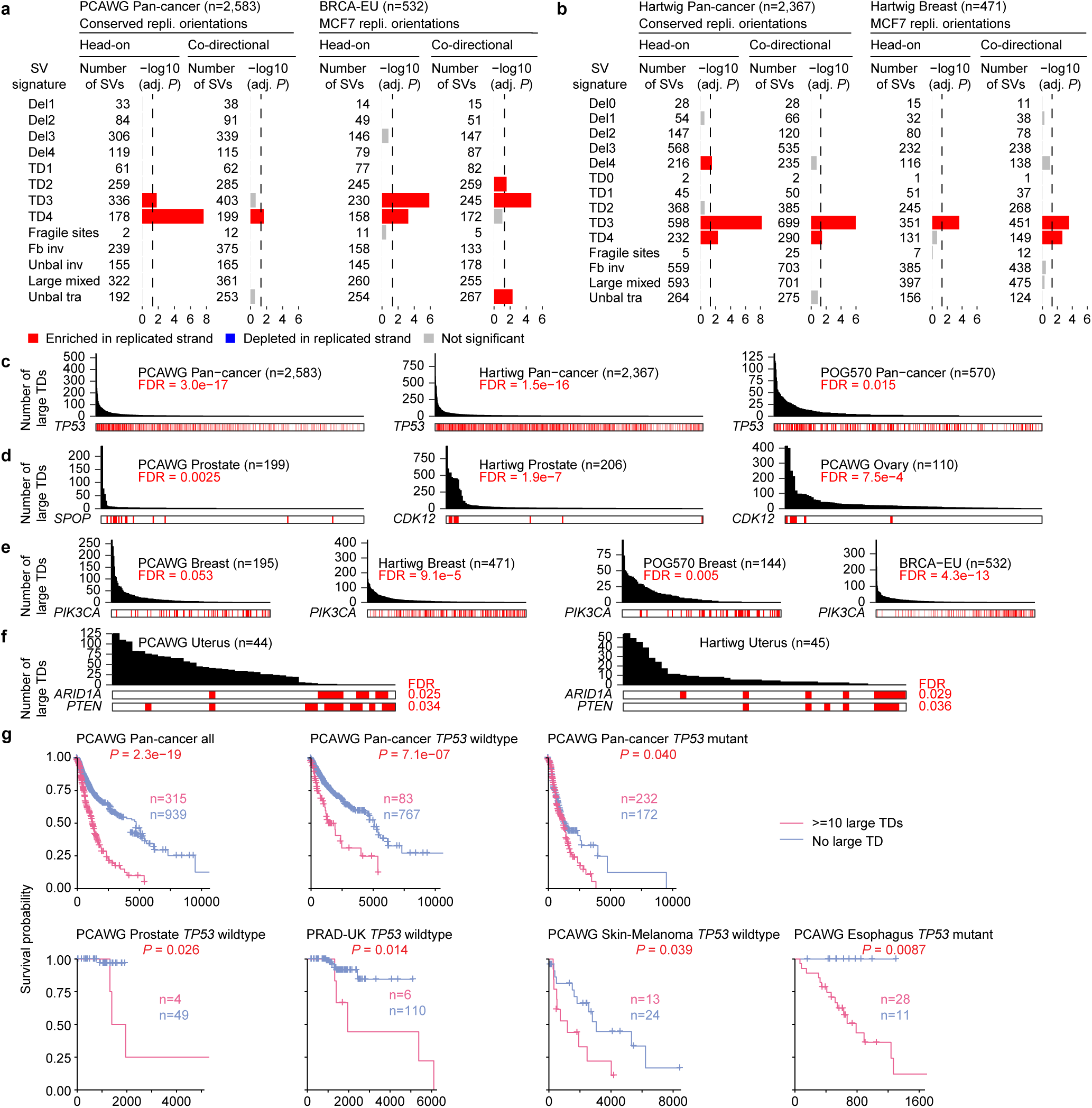
Replicated-strand bias in head-on and co-directional collision regions, and genetic and clinical associations of large TDs. **a** and **b**, Replicated-strand bias in head-on and co-directional collision regions in primary (**a**) and metastatic (**b**) tumors. The testing cohorts, tumor types, sample sizes, sources of replication orientation, head-on and co-directional collision regions are annotated on the top of the plots. *P* values are calculated by Fisher’s exact test. The x-axis shows Bonferroni-adjusted *P* values. **c**, *TP53* mutations positively associated with large TDs in the PCAWG, Hartwig, and POG570 cohorts. **d**, *SPOP* and *CDK12* mutations positively associated with large TDs in the PCAWG prostate, Hartwig prostate, and PCAWG ovarian cancers. **e**, *PIK3CA* mutations negatively associated with large TDs in four independent breast cancer cohorts. **f**, *ARID1A* and *PTEN* mutations negatively associated with large TDs in the PCAWG and Hartwig uterus cancers. For panels **c** to **f**, the x-axis denotes individual tumors. The red bars indicate tumors with protein-altering somatic mutations. Cohort names and sample sizes are annotated above the plots. *P* values are calculated by linear regression with mutation load as covariates. Benjamini-Hochberg procedure is used to derive false discovery rates (FDRs). **g**, Kaplan-Meier survival plots for patients with at least 10 large TDs and no large TD in their tumors. *P* values are calculated by the log-rank test. Cohorts and *TP53* mutation status are annotated on top of plots.

Many genomic features, such as GC content, transcription, replication, and chromatin state, are highly correlated. To rule out the possibility that replicated-strand bias in large TDs was due to other factors, we performed multivariate regression using SV strand (replicated or unreplicated) as the dependent variable and observed-VS-random SVs as well as a comprehensive list of genomic features as dependent variables. We found observed SVs were significantly associated with replicated strands only in TD3 and TD4 when other genomic features were controlled for (**Supplementary Fig. S11a**) in the PCAWG cohort. The same significant associations were also observed in the Hartwig cohort (**Supplementary Fig. S11b**) as well as the four breast cancer cohorts combined using replication timing from MCF7 (**Supplementary Fig. S11c**). Therefore, the replicated-strand bias in large TDs was an independent effect that could not be explained by other genomic features.

### Genetic and clinical associations of large TDs

We then sought to identify genes involved in TRC and large TD formation. We tested somatic SNVs in all protein-coding genes that altered coding sequences (**Supplementary Table S2**). As expected, *TP53* mutations were positively associated with large TDs (**Fig. 4c**) since they allow the cells to tolerate genome instability. *SPOP* mutations were also associated with large TDs in prostate cancer (**Fig. 4d**). Our recent finding that mutant SPOP is associated with complex SVs ^32^ suggests that mutant SPOP likely has a similar gatekeeper effect as mutant TP53. As we and others have previously reported ^40–43^, *CDK12* mutations, but not *BRCA1* or *BRCA2* mutations, are associated with large TDs in ovarian cancer and metastatic prostate cancer (**Fig. 4d** and **Supplementary Table S2**). *BRCA1* mutations are only associated with small TDs ^4,44^ but not large TDs. We focus on *CDK12* in greater detail in later sections. Interestingly, we found *PIK3CA* mutations were negatively associated with large TDs in four independent breast cancer cohorts (**Fig. 4e**). There are two possible reasons for somatic alterations to be mutually exclusive: the genes affect the same molecular pathway, or the alterations are incompatible (synthetic lethal). Although certain genes are recurrently altered by large TDs, large TDs affect many genes throughout tumor genomes. So, it is unlikely that large TDs always impact the same pathways as mutant PIK3CA. It is more likely that breast cancer cells with *PIK3CA* mutations cannot tolerate elevated TRCs. Moreover, *ARID1A* and *PTEN* mutations were negatively associated with large TDs in both primary and metastatic uterine cancers (**Fig. 4f**).

As mentioned above, large TDs recurrently amplify genes. In addition to genes previously reported, such as *MYC*, *MALAT1*, *MECOM*, *ESR1*, *ERBB2*, *FOXA1*, and *MDM2* ^37,45^, we found *GATA3* was frequently amplified in metastatic breast cancer, and *ERBB3* and *H3F3B* were frequently amplified in uterine cancer (**Supplementary Fig. S12**).

Next, we tested whether large TDs could be used for prognosis. Abundant large TDs in tumors were associated with poor overall survival when all tumor types were combined in the PCAWG cohort as well as when patients were stratified by tumor *TP53* mutation status (**Fig. 4g**). Abundant large TDs were also associated with poor survival in two prostate cancer cohorts (PCAWG and PRAD-UK), as well as melanoma and esophageal cancer (**Fig. 4g**).

### Loss of CDK12 leads to increased R-loops and TRCs

We have shown that *CDK12* mutations are associated with large TDs in cancer and that large TDs displayed footprints of TRCs. Therefore, we hypothesized that CDK12 inactivation causes elevated TRCs. To test this hypothesis, we generated single-cell derived CDK12 knockout (KO) clones using CRISPR/Cas9 in two prostate cancer cell lines—C42B and 22Rv1—and an immortalized normal prostate epithelial cell line—PNT2. A total of five CDK12 KO clones were generated (two in C42B, two in 22Rv1, and one in PNT2). CDK12 protein knockout was confirmed by western blot (**Fig. 5a**). In addition, we validated the knockouts by RNA-Seq to demonstrate that we had successfully generated bi-allelic indels in exon 1 of *CDK12*, leading to frameshifts and early truncations in both alleles (**Supplementary Fig. S13a**). This recapitulates the frameshift mutations found in primary prostate cancers and metastatic castration-resistant prostate cancers (CRPCs) (**Supplementary Fig. S13b** and **S13c**). While *CDK6* and *CDK14* were upregulated in C42B and 22Rv1 CDK12 KO clones (**Supplementary Fig. S14a**), the majority of other CDKs were largely unaffected (**Supplementary Fig. S14a** and **S15a**).

**Figure 5.**
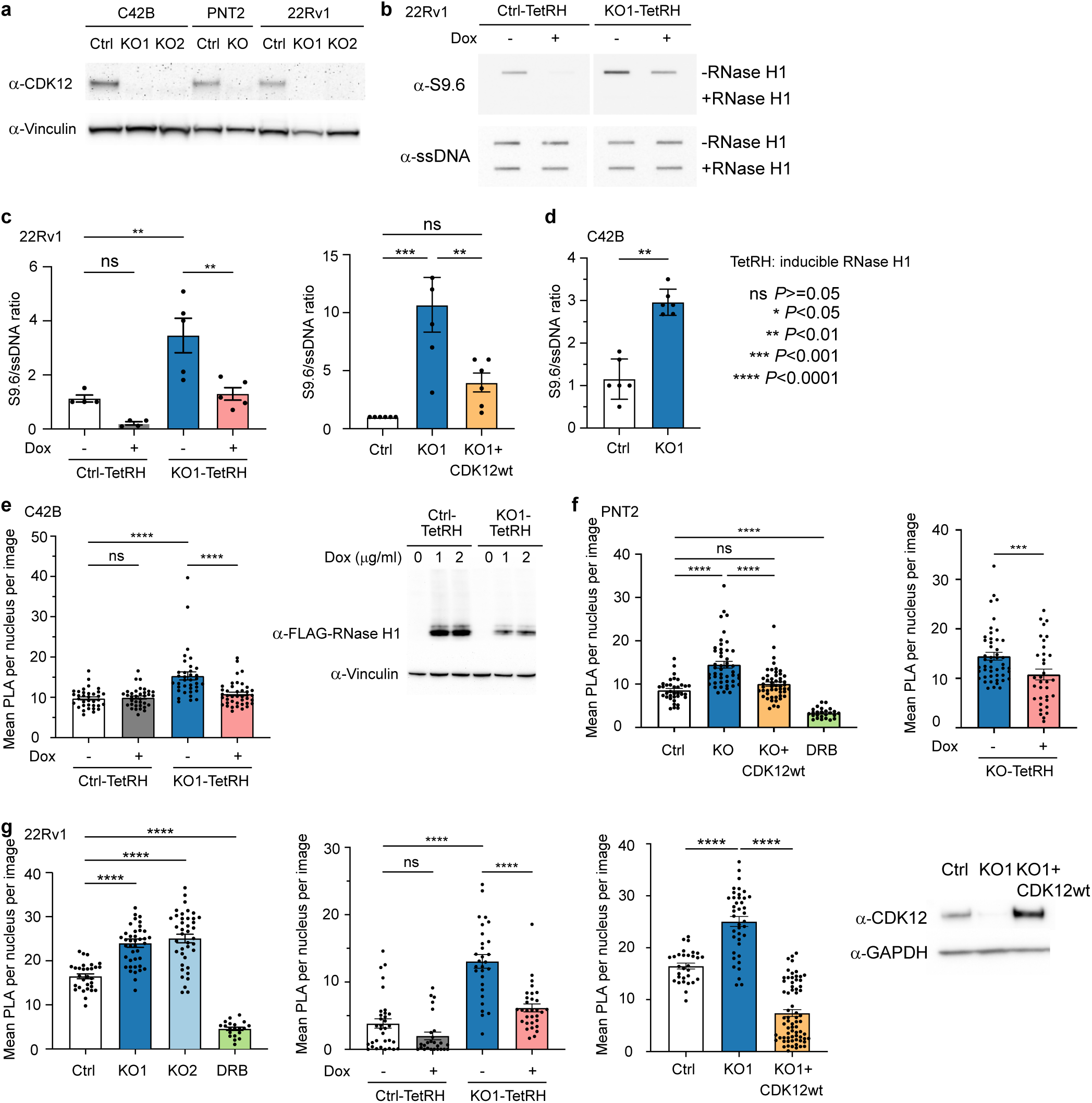
Loss of CDK12 results in increased R-loops and TRCs. **a**, Western blot (WB) showing CDK12 in control (Ctrl) and CDK12 KO cells in C42B, PNT2, and 22Rv1 cell lines. **b**, Slot blot in 22Rv1 Ctrl and CDK12 KO cells showing S9.6 (R-loops), with and without doxycycline-inducible RNase H1 (TetRH). RNase H1 addition is used as a control. Single strand (ss) DNA blot is shown as the loading control. **c**, Left panel: quantification of relative S9.6/ssDNA signal in 22Rv1 Ctrl and CDK12 KO1 cells (n=4 experiments in Ctrl cells and n=5 experiments in KO1 cells), with and without doxycycline-inducible RNase H1. Right panel: quantification of relative S9.6/ssDNA signal in 22Rv1 Ctrl, CDK12 KO, and CDK12 KO + CDK12wt re-expressing cells (n=6 experiments). **d**, Quantification of relative S9.6/ssDNA signal in C42B Ctrl and KO1 cells (n=6 experiments). **e**, Left panel: quantification of the mean PLA signal per image per nucleus in C42B Ctrl and KO1 cells (n=30 fields in one representative experiment), with and without doxycycline-induced expression of RNase H1. Right panel: WB showing doxycycline-induced expression of RNase H1, using FLAG antibody. **f**, Left panel: quantification of the mean PLA signal per nucleus per image in PNT2 Ctrl, CDK12 KO, and CDK12 KO + CDK12wt re-expressing cells (n=30 fields in one representative experiment). DRB (5,6-Dichlorobenzimidazole 1-β-D-ribofuranoside) treatment is used as a negative control. Right panel: Quantification of the mean PLA signal per nucleus per image in PNT2 CDK12 KO cells, with and without doxycycline-induced expression of RNase H1 (n=30 fields in one representative experiment). **g**, First panel: quantification of the mean PLA signal per nucleus per image in 22Rv1 Ctrl and CDK12 KO cells (KO1 and KO2) (n=30 fields in one representative experiment). DRB treatment is used as a negative control. Second panel: quantification of the mean PLA signal per nucleus per image in 22Rv1 Ctrl and CDK12 KO1 cells, with and without doxycycline-induced expression of RNase H1 (n=30 fields in one representative experiment). Third panel: quantification of the mean PLA signal per nucleus per image in 22Rv1 Ctrl, CDK12 KO1, and CDK12 KO1 + CDK12wt re-expressing cells (n=30 fields in one representative experiment). Fourth panel: WB showing CDK12 in 22Rv1 Ctrl, CDK12 KO1, and CDK12 KO1 + CDK12wt re-expressing cells. In **c**, **d**, **e**, **f**, and **g**, two independent experiments were performed. The *P* values are calculated using ANOVA and corrected for multiple tests. Boxplots show means (top bounds of boxes), standard errors of the mean (error bars), and individual data points (black dots).

CDK12 is known to control transcription through phosphorylation of RNA polymerase II (RNAPII) at serine 2 (Ser2) ^46,47^ and regulates transcription elongation ^48^ and termination ^49^. Upon CDK12 knockout, both total and chromatin fractions (protein bound to chromatin) of RNAPII Ser2 phosphorylation (pSer2) decreased slightly (**Supplementary Fig. S15b**) suggesting mildly impaired transcription. We then assessed the SV breakpoint locations relative to transcription start sites (TSSs) and transcription termination sites (TTSs) of protein-coding genes in both PCAWG and Hartwig cohorts. We found that large TDs, but not other types of SVs, were enriched near TTSs (**Supplementary Fig. S16**) which suggested that the TRC and large TD formation may be associated with impaired transcription termination and release of RNAPII. Next, we asked whether loss of CDK12 induces global changes in RNA-DNA hybrids, or R-loops. R-loops have previously been shown to be important regulators of genomic instability, particularly during DNA replication and DSB repair. Increased R-loops can impede efficient transcription and replication ^50–52^. We found that loss of CDK12 significantly increased total R-loops in 22Rv1 and C42B cells (**Fig. 5b**, **5c**, and **5d** “TetRH-Dox”). We next tested whether loss of CDK12 leads to increased TRCs, a potential consequence of increased R-loops that act as barriers to replication ^53^. To evaluate this, we performed a proximity ligation assay (PLA) to measure the colocalization of pSer2 RNAPII and the DNA replication protein, PCNA. We found that CDK12 KO cells had significantly increased PLA signals in all three cell lines, indicative of elevated TRCs (**Fig. 5e**, **5f**, and **5g** “TetRH-Dox”). The changes in R-loops and PLA could be rescued by re-expressing wildtype CDK12 in the CDK12 KO cells (**Fig. 5c**, **5f** and **5g**). Expression of RNase H1 in CDK12 KO cells reduced the R-loops and TRCs to levels comparable to CDK12 wildtype cells (**Fig. 5c**, **5e**, **5f, and 5g** “TetRH+Dox”). CDK12 KO cells also had elevated markers of replication stress (**Supplementary Fig. S15c-e**) including increased RPA32 phosphorylation and formation of RPA32 foci. To further test the functions of CDK12 using another silencing system, we generated CDK12 knockdown cells using CRISPR interference (CRISPRi) ^54^ and found similar increases in TRCs (**Supplementary Fig. S13d** and **S13e**). Taken together, these results demonstrated that loss of CDK12 induces R-loops and TRCs in both prostate cancer cells and normal prostate epithelial cells.

### Loss of CDK12 directly promotes large TD formation

To directly assess the effect of CDK12 loss on genome instability, we cultured the C42B cells (both KO and control) for a year (**Fig. 6a**) to allow enough time for somatic SVs to form. Since individual cells likely carried unique sets of SVs, we picked three single cells from both KO and control groups and expanded them into clones. WGS was performed on the six single-cell-expanded clones. The parental cells were sequenced at a much higher coverage (200x) so that pre-existing SVs, if there were any, could be removed. The control clones had 63 to 155 somatic SVs of all types, whereas the CDK12 KO clones had 40-78 SVs (**Fig. 6b**). Since C42B is a cancer cell line, and genome instability is a hallmark of cancer, it was expected that the control cells had ongoing genome instability and accumulated somatic SVs over the course of culturing. Control clone 3 had substantially more inversions and translocations than all other clones suggesting that there were other more active mechanisms capable of inducing somatic SVs in the cell during the one year in culture (**Fig. 6b**). Although different types of SVs and total number of SVs varied substantially among the six KO and control clones, the fractions of large TDs were less variable (**Fig. 6c**). The CDK12 KO clones had significantly more large TDs (15.2% on average) than control clones (4.7% on average) (**Fig. 6c**). These results indicate that loss of CDK12 can directly promote large TD formation.

**Figure 6.**
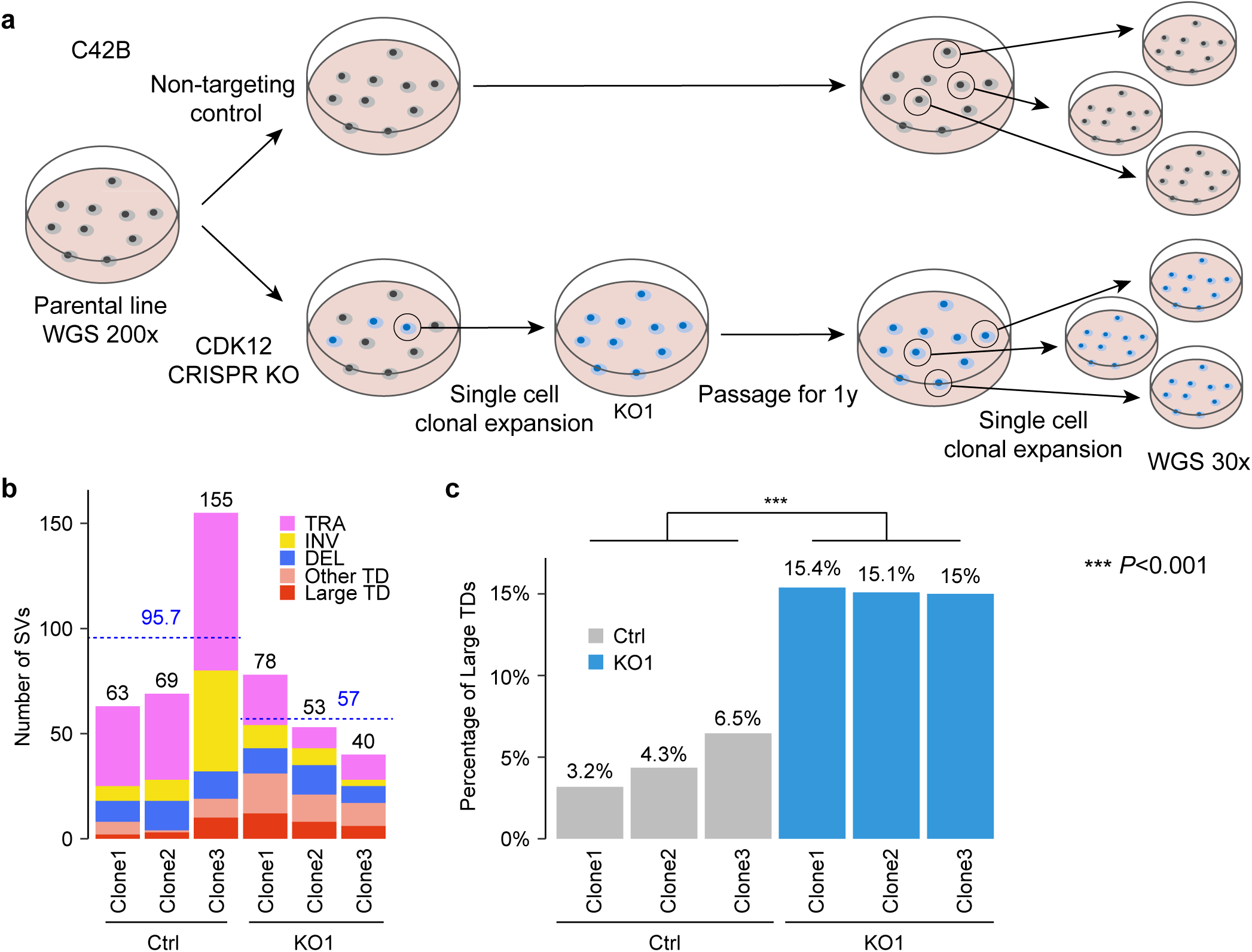
Loss of CDK12 promotes large TD formation. **a**, Workflow for single cell clonal expansion in C42B cells. **b**, Number of somatic SVs in three Ctrl clones and three CDK12 KO1 clones. The total numbers of somatic SVs are indicated by black numbers above the bars. The dashed blue lines and blue numbers indicate the average numbers of somatic SVs. **c**, The percentages of large TDs in three Ctrl clones and three CDK12 KO1 clones. The *P* value is calculated using Student’s t test.

### Large TDs as a biomarker for drug sensitivity

Since cells lacking CDK12 had elevated TRCs, we hypothesized that they may be more dependent on certain cellular pathways that can be targeted by drugs. Based on RNA-Seq data, the G2/M checkpoint was up-regulated in CDK12 KO cells for both C42B and 22Rv1 (**Supplementary Fig. S14b**). In addition, there were fewer cells in G2/M when CDK12 was inactivated (**Supplementary Fig. S17**). These results suggested that cells without CDK12 may be more dependent on the G2/M checkpoint. We then performed an unbiased in silico drug screen to assess drug sensitivity using large TDs as a biomarker. We used the 325 cell lines in the Cancer Cell Line Encyclopedia (CCLE) that have WGS data, allowing us to accurately quantify large TDs. Among these cell lines, large-scale experimental drug screening ^55^ has been conducted in 203 cell lines. We focused on drugs related to DNA damage, damage repair, DNA replication, and transcription (**Supplementary Table S3**), and found that cancer cell lines with abundant large TDs were significantly more sensitive to two reagents that inhibit DNA repair— the WEE1 inhibitor MK-1775 (also known as AZD-1775 or adavosertib) and the PARP inhibitor PJ-34 (**Fig. 7a**). Interestingly, WEE1 is a nuclear kinase regulating G2/M checkpoint transition and allows DNA repair before mitotic entry ^56^. MK-1775 has shown potent activities when combined with DNA damaging agents ^57^. We found that in all three cell lines, loss of CDK12 significantly increased drug sensitivity to MK-1775 as well as other WEE1 inhibitors, such as Debio 0123, ZN-c3, and PD0166285 (**Fig. 7b**, **7c**, **7d** and **Supplementary Fig. S18a-c**). CDK12 KO cells treated with MK-1775 also had more DSBs formed, consistent with elevated DNA damage (**Supplementary Fig. S18d**). Although phase II clinical trials of MK-1775 have been conducted in ovarian, breast, lung, gastric cancers, etc. ^58^, our results suggest that patients with other tumor types, such as prostate, esophageal cancers, and even a small subset of liver cancers with high frequencies of large TDs (**Fig. 2d** and **2e**), may benefit from WEE1 inhibition. Moreover, because CHK1 and ATR are upstream of WEE1 in G2/M checkpoint pathway activation ^56^, we further tested CHK1 and ATR inhibitors and found that CDK12 KO cells were significantly more sensitive to multiple CHK1 and ATR inhibitors including prexasertib, CHIR-124, AZD7762, CCT245737, ceralasertib, VX-970, and VE-821 (**Fig. 7d** and **Supplementary Fig. S18a-c**). Finally, we found that cancer cell lines abundant in large TDs were resistant to three DNA synthesis inhibitors—bendamustine (DNA alkylating agent), nelarabine (guanine analog) and famciclovir (another guanine analog) (**Fig. 7a**). It is possible that cancer cells with abundant large TDs have replication stress due to frequent TRCs and are already impaired in DNA synthesis. Therefore, adding additional replication stress by blocking DNA synthesis in these cells does not further enhance cell killing. Taken together, our results demonstrate that targeting the G2/M checkpoint may be a therapeutic strategy in tumors deficient in CDK12 and other cancers characterized by abundant large TDs.

**Figure 7.**
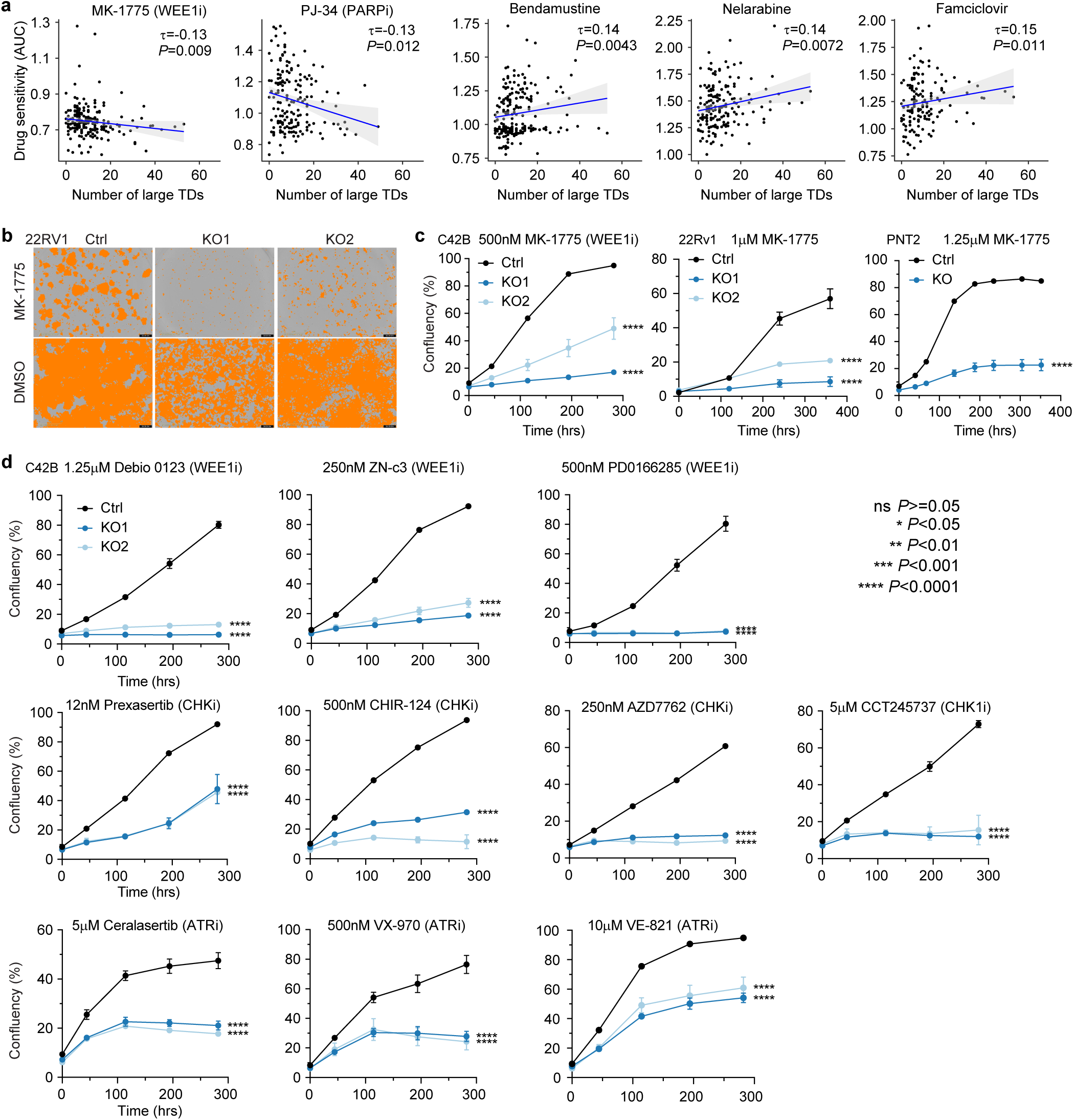
Drug sensitivity. **a**, Drug sensitivity measured by area under the curve (AUC) in association with the number of large TDs in cancer cell lines. Smaller values of AUC indicate that cells are more sensitive to the drug, whereas large values indicate that cells are more resistant. *P* values are calculated by Kendall test. **b**, Phase contrast images shown with the confluency mask (orange) for 22Rv1 Ctrl and CDK12 KO cells (KO1 and KO2) treated with DMSO or 1 μM of MK-1775 from day 15. **c**, Growth curves of Ctrl and CDK12 KO cells treated with the indicated doses of MK-1775 in C42B, 22Rv1, and PNT2 cell lines. **d**, Growth curves of C42B Ctrl and CDK12 KO cells (KO1 and KO2) treated with the indicated doses of three WEE1 inhibitors (Debio 0123, ZN-c3, and PD0166285), four CHK inhibitors (prexasertib, CHIR-124, AZD7762, and CCT245737), and three ATR inhibitors (ceralasertib, VX-970, and VE-821) for indicated times. In **c** and **d**, error bars represent standard errors of the mean. The *P* values are calculated by Student’s t test at the final timepoint for each experiment. Significant levels are indicated by “*” next to the corresponding KO cells compared to Ctrl cells.

### Proposed model

We propose the following model to explain our observations (**Fig. 8**). In normal cells, CDK12 phosphorylates RNAPII. In tumors with complete loss of CDK12, RNAPII is not properly phosphorylated, which impairs transcription, including the termination process. R-loops are formed when RNAPII cannot be properly released from the DNA template, which subsequently blocks the DNA polymerase complex and replication fork. Once TRCs occur, most of them are properly repaired. However, only in rare instances (approximately once per month per cell), the stalled replication forks are not properly repaired and collapse. The resulting chromosomal breaks generate DNA DSBs and eventually lead to large TDs.

**Figure 8.**
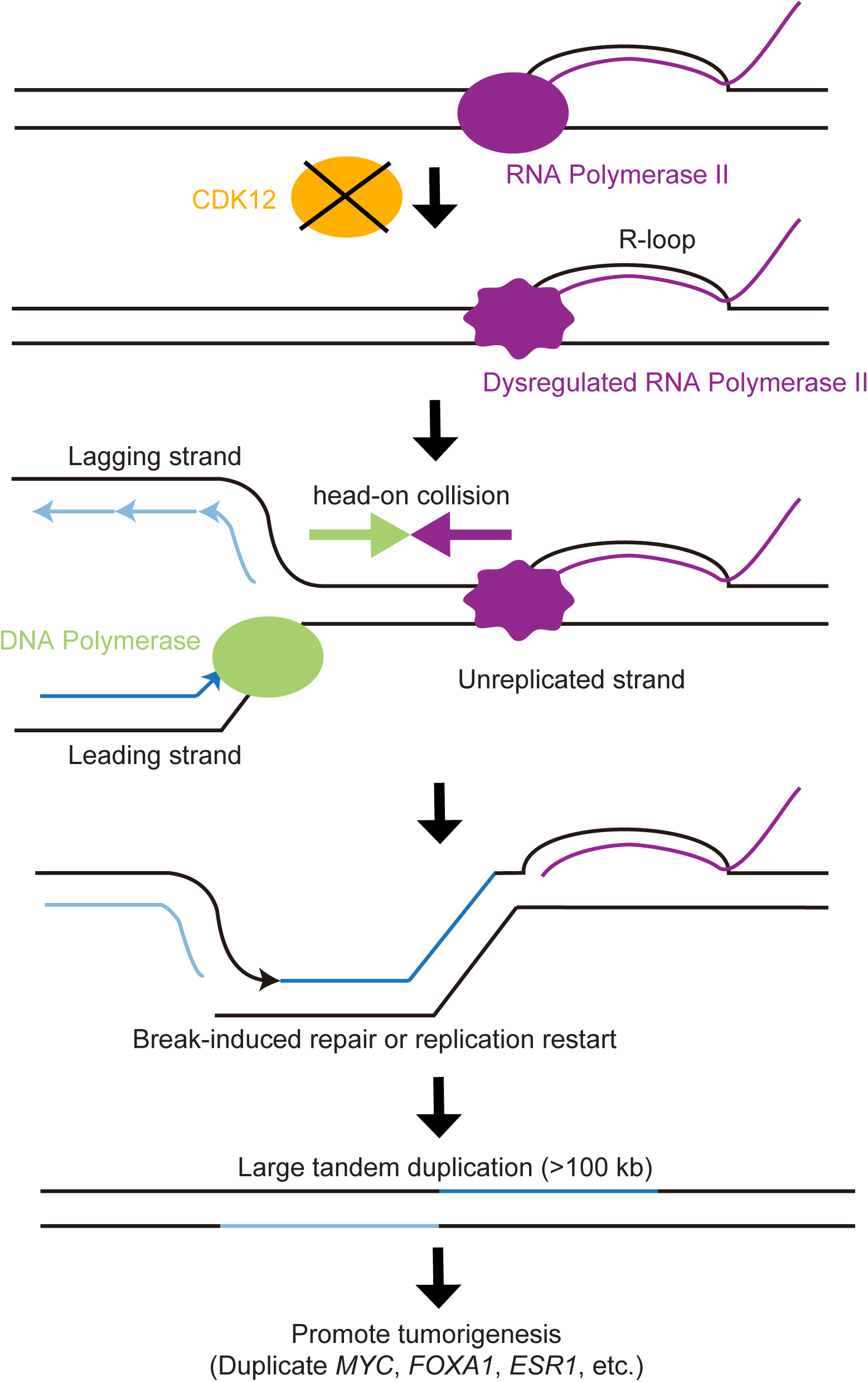
Final model. Working model of CDK12 loss leading to increased R-loops, TRCs, and large TDs.

There remain multiple open questions, such as how TRCs lead to large TDs. The DNA damage repair (DDR) system certainly plays an important role in SV formation, likely recognizing replication fork blockage, DSBs, and stretches of single strand DNA (ssDNA), and activating cell cycle checkpoints mediated by ATM and ATR ^59^. In addition, BRCA1 and BRCA2 have been shown to play roles in resolving R-loops ^60–62^. In the context of CDK12 deficiency, prior studies have shown that CDK12 controls the expression of many DNA repair genes, including BRCA1 and ATM, by regulating intronic polyadenylation ^63–65^. The global DDR deficiency resulting from CDK12 loss may lead to aberrant repair of TRCs. Therefore, it is also possible that loss of CDK12 not only directly promotes the formation of TRCs, but also leads to TRC repair deficiency which subsequently causes the accumulation of TRCs in CDK12 deficient cells.

Another open question is why TRCs specifically generate large TDs, but not other types of SVs, such as deletions and translocations. The types of SVs generated from DSBs depends on the repair partner. For example, translocations will form if the broken DNA ends are ligated to other chromosomes. TDs being produced by TRCs suggests that the collapsed replication forks fold backwards and are joined with repair partners upstream of the collision sites. A previous study showed that stalled replication forks lead to small TDs in mutant BRCA1 background via three possible mechanisms—breakage fusion, microhomology-mediated break-induced repair (MMBIR), or replication restart-bypass ^44^. The partner sister strand is rearranged in the breakage-fusion mechanism, whereas it is unrearranged in the latter two mechanisms. In cancer, large TDs do not co-occur with large deletions at the same loci, suggesting that large TDs likely form through either MMBIR or replication restart-bypass (**Fig. 8**). From an evolutionary point of view, forming a large TD is probably the optimal solution for cells not able to restart replication at the collapsed forks after the collision. Since TRCs always occur in genes, at each collision site, part of the gene is replicated and the rest of the gene is unreplicated. Forming other types of SVs, such as deletions, inversions, and translocations, will lead to failure to replicate the genes at the collision sites which is likely more deleterious to the cell. The only way to preserve the gene sequences is to re-replicate these regions and thus, form TDs. If the replication forks fold backwards close to the collision sites and form small TDs, DNA polymerase complexes will reach the collision sites rapidly. If the collisions have not yet been cleared, more collisions will occur. Thus, an optimal way to avoid additional collisions is to fold backwards far away from the collision sites, hundreds of kb upstream, so that when the DNA polymerase complexes reach the collision sites, the RNAPIIs are released, and the R-loops are resolved. Therefore, large TDs are formed.

Although previous work showed that co-directional collision reduces R-loop formation ^18^, we found no significant differences in large TDs between head-on and co-direction collisions except in the PCAWG cohort (**Fig. 4a** and **4b**). Further mechanistic studies are required to fully understand the formation of R-loops, TRCs, and large TDs. Nonetheless, our data suggest that R-loops, TRCs, and large TDs are promoted by CDK12 loss.

Finally, a subset of large TDs result in duplications of numerous well-known oncogenes; therefore, some large TDs are under positive selection and may contribute to tumorigenesis (**Fig. 8**).

## Discussion

Here, we report that large TDs in tumors result from accumulation and improper repair of TRCs. We first deconvoluted highly conserved somatic simple SV signatures from three independent pan-cancer WGS cohorts (the PCAWG, Hartwig, and POG570). We reasoned that SVs formed via TRC would display replicated-strand bias. Indeed, we observed such bias in two large TD signatures. The replicated-strand bias in large TDs depended on transcription, could not be explained by any other genomic factors, and was reproducible in two pan-cancer cohorts as well as several other independent cohorts. Furthermore, we observed that *CDK12* mutations were associated with abundant large TDs in prostate and ovarian cancers. Experimentally, we demonstrated that knocking out CDK12, a key regulator of mRNA transcription ^66^, led to elevated R-loops, TRCs, and large TDs.

The replication orientations were inferred in a manner similar to a previous study on mutational strand asymmetries in cancer ^67^. The “left-replicated” and “right-replicated” regions were defined as “sharp transition” regions and the “flat” regions (uniformly early- or uniformly late-replicated) were ignored. We only used sharp transition regions to ensure high confidence of inferred replication orientations. Replication orientations in flat regions cannot be clearly defined and SV strand bias cannot be tested. Therefore, these regions were not used in this study. However, TRCs may still occur in flat regions. In tumors, large TDs being most enriched in the earliest replicated regions (**Supplementary Fig. S5** highest peaks in left most regions for TD3 and TD4) suggested that TRCs occur frequently in early replicated regions. This is consistent with early replicated regions being enriched in genes that are more transcriptionally active which leads to frequent TRCs.

Large TDs are abundant in several tumor types, such as uterine, breast, ovarian, prostate, esophageal, stomach, and head and neck cancers **(Fig. 2d** and **2e**). Most tumors with abundant large TDs are female cancers and upper gastrointestinal tract cancers, which suggests that the tissues of origin may play a role. Oncogene activation is known to cause replication stress ^12–17^. However, we did not observe activating mutations in oncogenes, such as *BRAF*, *KRAS*, *EGFR*, and *PIK3CA*, to be positively associated with large TDs in any cohorts. Instead, *PIK3CA* mutations were negatively associated with large TDs in four independent breast cancer cohorts (**Fig. 4e**). These results suggest that large TDs may not arise directly from oncogene-induced replication stress.

Selectively targeting cancers harboring large TDs would be of considerable potential therapeutic utility. Prior studies suggested that large TDs may be a source of neoepitopes resulting from gene fusions, which can be recognized by the immune system ^41^. A phase II clinical trial evaluating checkpoint immunotherapy in prostate and other cancers with *CDK12* mutations is currently underway ^68^. Given the possible deficiencies in TRC repair and DNA replication in tumors with large TDs, we sought to uncover synthetic lethal vulnerabilities caused by CDK12 loss. Our data demonstrated that multiple G2/M checkpoint proteins, such as WEE1, CHK1, and ATR, may be important therapeutic targets in this context, and that large TDs and *CDK12* mutations may be attractive biomarkers for drug sensitivity.

## Data availability

The patient cohorts used in this study are summarized in **Supplementary Table S1**. There were 2,583 primary tumors in the PCAWG cohort ^33^, 2,367 metastatic tumors in the Hartwig Medical Foundation cohort ^34^, 570 metastatic tumors in the POG570 cohort ^35^, 532 primary breast cancers (BRCA-EU), 115 primary ovarian cancers (OV-AU) and 218 primary prostate cancers (PRAD-UK). Somatic variants (SNVs, CNVs and SVs), gene expression quantifications and clinical information (tumor histology and patient survival) were obtained from https://dcc.icgc.org/pcawg for PCAWG cohort, https://hartwigmedical.github.io for Hartwig cohort, https://www.bcgsc.ca/downloads/POG570/ for POG570 cohort, https://dcc.icgc.org/ for OV-AU and PRAD-UK cohorts. Gene expression data of BRCA-EU were provided by Dr. Marcel Smid. A subset of breast cancers in the PCAWG cohort had molecular subtypes annotated in a previous study ^69^. Gene expression quantifications from tumor adjacent normal samples of the Cancer Genome Atlas (TCGA) cohort were downloaded from Genomic Data Commons (https://portal.gdc.cancer.gov/). Somatic SVs (CCLE2019 release) in 328 cancer cell lines and drug response data (PRISM Repurposing 19Q4) ^55^ were downloaded from the Cancer Dependency Portal (https://depmap.org/portal/download/all/). Human reference genome assembly GRCh37 (hg19) was used in the entire study. Reference genome mappability was obtained from https://bismap.hoffmanlab.org/.

The raw replication timing data for fourteen cell lines derived from solid tissues (A13, BG01 NPC, BG02 ESC, CyT49_Definitive_Endoderm, CyT49_Liver, CyT49_Neural_Crest, CyT49_Panc, HCT116, HEK293, HeLaS3, HepG2, MCF7, RPE1 hTERT, and U2Os) were downloaded from https://www2.replicationdomain.com. The replication timing data of GM12878, IMR90, and NHEK were obtained from a previous study ^70^.

Oncogenes were obtained from the COSMIC cancer gene census (https://cancer.sanger.ac.uk/census). GC content was calculated using bedtools for 50bp windows flanking both sides of each SV breakpoint. CpG islands, centromere, telomere, repeat annotation, and Lamina associated domains (LAD) for human Tig3 lung fibroblasts were downloaded from UCSC Genome Table Browser (https://genome.ucsc.edu/). Non-B DNA annotation was downloaded from non-B DB ^71^ (https://nonb-abcc.ncifcrf.gov/). ENCODE Chromatin Immunoprecipitation Sequencing (ChIP-Seq) data for H3k27ac, H3k27me3, H3k36me3, H3k4me1, H3k4me2, H3k4me3, H3k79me2, H3k9ac, H3k9me3, H4k20me1 of GM12878 cell line were downloaded from the UCSC composite track (http://genome.ucsc.edu/cgi-bin/hgFileUi?db=hg19&g=wgEncodeUwHistone). Nucleosome occupancy for the K562 cell line was downloaded from ENCODE (https://www.encodeproject.org/files/ENCFF000VNN/). Topologically associating domain (TAD) boundaries of GM12878 ^72^ were downloaded from NCBI (https://www.ncbi.nlm.nih.gov/geo/query/acc.cgi?acc=GSE63525).

Raw sequencing data of WGS and RNA-Seq for CDK12 KO and control clones are deposited in the National Institutes of Health (NIH) Sequence Read Archive (SRA) under BioProject ID PRJNA1055686.

All data described above are publicly available.

## Code availability

Custom scripts used in this study can be found at: https://github.com/yanglab-computationalgenomics/TRC.SV.strand.bias

## Methods

### Simple SVs and simple SV signatures

Clustered complex SVs detected by Starfish and non-clustered complex SVs, such as chromoplexy and cycles of insertions, detected by junction patterns ^2^ were removed from raw somatic SVs. The remainder of SVs were simple SVs and classified into nine types—deletions, fragile site deletions, TDs, fragile site TDs, foldback inversions (breakpoints within 50kb in distance), unbalanced inversions, reciprocal inversions, unbalanced translocations, and reciprocal translocations (**Supplementary Fig. S2**). Fragile site annotation was obtained from the PCAWG SV study ^2^. Deletions, TDs, and inversions were further divided by size (**Fig. 1b**). For deletions and TDs, we divided each magnitude of size range into 4 categories (i.e., 10 kb - 100 kb was divided into 10 kb - 25 kb, 25 kb - 50 kb, 50 kb - 75 kb, 75 kb - 100 kb). With the two boundary categories (0 - 1 kb and >10 Mb), there were a total of 18 categories (**Fig. 1b** and **1c**). As a result, a total of 49 simple SV classes were defined (**Fig. 1b**). Simple SV signatures were extracted using SigProfilerExtractor ^36^ with default parameters in the PCAWG, Hartwig, and POG570 cohorts. In the PCAWG cohort, deletions ranging in size 0 - 5 kb, 5 kb - 25 kb, 25 kb - 500 kb, 500 kb - 5 Mb were assigned to signatures “Del1”, “Del2”, “Del3”, and “Del4”, while TDs ranging in 0 - 10 kb, 10 kb - 100 kb, 100 kb - 500 kb, 500 kb - 7.5 Mb were assigned to signatures “TD1”, “TD2”, “TD3”, and “TD4”. Deletions greater than 5 Mb, TDs greater than 7.5 Mb, reciprocal inversions, and reciprocal translocations were assigned to signature “Large mixed”. In the Hartwig cohort, deletions ranging in size 0 - 1 kb, 1 kb - 5 kb, 5 kb - 25 kb, 25 kb - 500 kb, and 500 kb - 5 Mb were assigned to signatures “Del0” to “Del4”, TDs ranging in 0 - 1 kb, 2 kb - 10 kb, 10 kb - 100 kb, 100 kb - 750 kb, and 750 kb - 7.5 Mb were assigned to signatures “TD0” to “TD4”. Foldback inversions and unbalanced inversions less than 5 Mb were assigned to signature “Fb inv”. Deletions greater than 5 Mb, TDs greater than 7.5 Mb, unbalanced inversions greater than 5Mb, reciprocal inversions, and reciprocal translocations were designated to signature “Large mixed”.

### Replication orientation annotation

For each somatic SV observed in tumors, two random SVs were generated by placing the same SV in random uniquely mappable locations of the same chromosome with SV type and size preserved. Replication timing data were re-scaled to a range between 0 and 1 and were annotated for observed and random SV breakpoints. A two-sided Kolmogorov-Smirnov test was employed to compare observed breakpoints and random breakpoints. Benjamini-Hochberg procedure was used to calculate False Discovery Rates (FDRs). The procedure of testing replication timing was exactly the same as the PCAWG SV signature study ^2^.

The replication timing data were quantile normalized across all cell lines. The reference genome was divided into bins of 50 kb in size and the normalized values were averaged within the bins. The replication timing slope was calculated for adjacent bins, and then the replication orientations, “left-replicated” and “right-replicated”, were determined for sharp transition bins (slope < 5e-7) but not for flat regions (**Supplementary Fig. S8**). The replication orientations of protein-coding genes were assigned accordingly if the genes were entirely located in either left-replicated or right-replicated regions. There were between 6,581 to 9,548 genes annotated as left- and right-replicated per cell line. If the replication orientations of a gene were the same in at least 8 out of 14 cell lines, the gene was considered to have conserved replication orientation. There were 3,493 genes with conserved replication orientations in solid tissues. Protein-coding genes with the opposite transcription and DNA replication orientations would have head-on collisions whereas the ones with the same transcription and DNA replication orientations would have co-directional collisions.

### Strand bias test

SV breakpoint orientations were assigned as in **Supplementary Fig. S7.** Deletions and TDs entirely residing within protein-coding genes (both breakpoints located in the same genes) were discarded because deletions and TDs have both “+” and “-” breakpoints (**Supplementary Fig. S7**). We reasoned that even if they truly resulted from transcription and replication collision, the strand effect would be cancelled out by the mixture of “+” and “-” breakpoints in these SVs. The breakpoints of the remainder of SVs were considered individually. Only the breakpoints located in non-overlapping protein-coding genes with defined replication orientations were used to test for strand bias. SV breakpoints located in overlapping protein-coding genes were discarded because these gene often had different expression levels. Breakpoints in intergenic regions or in genes without defined replication orientations were also discarded. Then, each SV breakpoint in protein-coding genes was assigned as “replicated” or “unreplicated” based on the breakpoint orientation and replication orientation. For each SV signature, a two-by-two table (“replicated/unreplicated” and “observed/random SVs”) was constructed and replicated-strand bias in observed SVs was tested by two-sided Fisher’s exact test followed by Bonferroni correction. Corrected *P* values that were less than 0.05 were considered significant. All protein-coding genes were ranked based on the expression levels (upper-quartile normalized FPKM or adjusted TPM) in each tumor sample when RNA-Seq data were available, and the top 75% genes were defined as “expressed” whereas the bottom 25% were “unexpressed”. When defining “expressed” and “unexpressed” genes by gene expression in normal samples, genes were ranked by median expression levels of adjacent normal samples in tumor types from TCGA of which RNA-Seq data were available for at least 20 normal samples.

Logistic regression was performed to take other genomic features into account in addition to gene expression and replication timing. The genomic features included GC content, CpG island, SV breakpoint distance to centromere, breakpoint distance to telomere, breakpoint distance to various repetitive elements (Alu, L1, L2, LTR, MIR elements, DNA transposons and low complexity repeats), breakpoint distance to non-B DNA motifs, histone markers, nucleosome occupancy, LAD, and breakpoint distance to TAD boundaries. All distances were log10 transformed. Signal enrichment values (density of modifications in 25 bp window) of histone modifications at the SV breakpoints were used. The proportion of bases in LAD at a 1 Mb window flanking breakpoints were calculated. The dependent variable was replicated/unreplicated strand of SV breakpoints. The above features, along with observed/random SV, gene expression, replication timing, and the interaction of gene expression and replication timing, were independent variables.

### Genetic and clinical associations of large TDs

Somatic SVs in TD3 and TD4 in both PCAWG and Hartwig cohorts were defined as large TDs (TDs with sizes ranging between 100 kb and 7.5 Mb not in fragile sites). Linear regression was performed to test the associations between frequencies of large TDs and genes with protein-coding altering SNVs. Mutation load was included as a covariate in all regression models and tumor type was modelled as a covariate in the pan-cancer analysis. Large TD frequency and mutation load were log10 transformed. Only protein-coding genes with mutation frequencies of at least 3% at pan-cancer level and at least 4% in individual tumor types were considered. FDR was calculated by Benjamini-Hochberg procedure, and 0.1 was used as the significance cutoff. A 10% mutation frequency cutoff was used for uterus cancers in the Hartwig cohort due to their high mutation load. GISTIC2 ^73^ was used to identify genomic regions with recurrent large TDs. The confidence level was set at 0.9 and default values were used for all other parameters. Log rank test was performed for survival analysis. Patients with at least 10 large TDs in their tumors were compared to those without any large TDs stratified by *TP53* mutation status and tumor type. The full length of protein-coding genes from transcription start sites to transcription termination sites was split into four quarters. All SV breakpoints were mapped to these quarters. A two-sided Fisher’s exact test was used to calculate the enrichment of observed breakpoints over random breakpoints within a specific quarter compared to other quarters. Bonferroni correction was performed on *P* values.

### In silico drug sensitivity test

Complex SVs in CCLE cell lines were removed similar to patient tumors. Three cell lines, LK2_LUNG, SNU16_STOMACH, and NCIH1436_LUNG, were discarded due to the presence of too many likely artifactual small TDs. Only drug dose response scores from the HTS002 screen were used ^55^ since it was the largest screen (1,394 drugs, 480 cell lines and 592,912 drug x cell line combinations). Drugs in the following categories were tested—DNA damage agents, DNA damage repair inhibitors, DNA replication inhibitors, and transcription inhibitors (**Supplementary Table S3**). For each drug, Kendall’s Tau correlation coefficient was calculated between large TD frequencies and areas under the ROC Curve (AUCs) of the drug in all cell lines.

### Cell culture

22Rv1 cells were obtained from the American Type Culture Collection (ATCC), C42B cells were obtained from the University of Michigan, and PNT2 cells were obtained from Sigma. Cells were grown in standard conditions with antibiotics (RPMI-1640 with 10% FBS and penicillin/streptomycin) and routinely tested for mycoplasma (Lonza). Cell lines were all validated by STR DNA profiling (Genetica). Cells were transduced with NucLightRed lentivirus (Sartorius) according to the manufacturer’s instructions. For doxycycline-induction, cells were treated with 1-2 μg/ml of doxycycline (Sigma) for 48 hours.

### Plasmids and cloning

pSpCas9(BB)-2A-GFP (PX458) (Addgene plasmid # 48138) was obtained from Dr. Feng Zhang. pDONR221 was from Thermo Fisher Scientific. Full-length human CDK12 cDNA was a gift from Dr. Gregg Morin and Dr. Grace Cheng as previously described ^74^, and then cloned into pDONR221 and pLVX-CMV-V5-Blast. Full-length human RNase H1 was obtained from Addgene (Addgene plasmid #65783), and then cloned into the pDONR221 and pLV-TetON-FLAG-puro as previously described ^75^. For Gateway cloning, BP and LR Clonase II (Thermo Fisher Scientific) reactions were performed according to the manufacturer’s protocol. Cloning was performed using standard protocols. For CRISPR interference (CRISPRi) assays, cells were transduced with dCas9 from pHR-SFFV-dCas9-BFP-KRAB (Addgene plasmid #46911) and sgRNA sequences were cloned into pCRISPRia-v2 (Addgene plasmid #84832), both from Dr. Jonathan Weissman. Gel purification was performed using the NucleoSpin Gel and PCR Clean-up kit (Takara Bio Inc.) according to the manufacturer’s protocol. Annealed sgRNAs were ligated into the plasmid using the Quick Ligation Kit (New England BioLabs) according to the manufacturer’s protocol.

### Lentiviral production

Lentiviral production was carried out using calcium-phosphate-mediated transfection of HEK293T-Lenti-X cells using standard procedures. Lentivirus was concentrated using the Lenti-X concentrator (Takara Bio Inc.) according to the manufacturer’s protocol. Stably transduced cells were selected with blasticidin (3 μg/ml, Gibco) or puromycin (3 μg/ml, Gibco) for at least 5 days.

### Generation of CDK12 knockout and knockdown cells

C42B, 22RV1 and PNT2 cells were transfected with the PX458 plasmid containing the CRISPR/Cas9 and the guide targeting CDK12 using Lipofectamine 3000 (Thermo Fisher) according to manufacturer’s instructions. 48-72 hours after transfection, cells were sorted for green fluorescence protein (GFP) expression, and then into single cells in a 96-well plate format using a FACS Aria. Guide sequences to knockout CDK12 were sg1: 5’ GCTTGTGCTTCGATACCAA 3’ and sg2: 5’ ACTTTGCAGCCGTCATCGGG. Clones were expanded and validated by western blot and by sequencing the target site. Non-targeting guide RNA was used as negative control. To knockdown CDK12 using CRISPR interference (CRISPRi) the following guide was used: 5’ GTGAGAAGCCGACGGCCCG 3’ (CDK12sg).

### Somatic SVs in C42B cells with CDK12 loss

The single-cell expanded clones were whole-genome sequenced at 30x coverage and the parental C42B cells were sequenced at 200x coverage by Illumina NovaSeq X Plus at Novogene. Reads were aligned by BWA mem 0.7.17 ^76^. Somatic SVs in single-cell expanded clones were called using Meerkat ^1^, Manta ^77^, and Delly ^78^. SVs identified by Manta required at least five supporting read pairs and one split read. Intra-chromosomal SVs called by Delly that were smaller than 500 bp in size required at least four supporting read pairs and one split read. SVs detected by different algorithms were merged when their breakpoints were on the same chromosomes, with the same orientations, and were within 10 bp. SVs detected by the three algorithms were combined and then filtered by the parental cell line and unrelated clones using the Meerkat filtering processes to ensure that pre-existing SVs in the parental cells were removed. All somatic large TDs were manually inspected in genome browser to ensure high confidence.

### RNA-Seq and gene expression analysis

RNA was extracted from cells using the Quick RNA Miniprep Kit (Zymo Research). RNA-Seq was performed by Illumina NovaSeq at Novogene. After removing low quality reads, each sample yielded about 50 million reads. Reads were aligned with STAR 2.7.10b. Differentially expressed genes were identified by DESeq2 ^79^ and gene set enrichment analysis was performed by GSEA 4.3.2 ^80^.

### Western blot

For total protein lysates, cells were lysed in RIPA buffer the Halt protease and a phosphatase inhibitor cocktail (Thermo Fisher Scientific). For chromatin fractionations, freshly collected cells were processed using the ChromaFlash Chromatin Extraction Kit according to manufacturer’s instructions (EpigenTek). Cells were treated with 2mM hydroxyurea (MedChemExpress) for 48 hours prior to cell collection and lysis. Protein concentration was measured using the BCA Protein Assay Kit (Thermo Scientific). Lysates were mixed at a 1:1 ratio with Laemmli Buffer (Bio-Rad) supplemented with beta-mercaptoethanol, boiled at 95°C, and then subjected to SDS-PAGE, transferred to PVDF membranes using the Trans-Blot Turbo transfer system (Biorad), blocked in 5% w/v BSA, incubated with primary antibody overnight. The next day, Horseradish Peroxidase (HRP) conjugated secondary antibodies (Cell Signaling, #7074, #7076 and #7077, 1:8000) were used and the blot was visualized using ECL Detection Reagents (Genesee Scientific). Primary antibodies used included: CDK12 (Atlas #HPA008038, 1:1000), GAPDH (Cell Signaling, #2118, 1:2000) Lamin B1 (Cell Signaling, #13435, 1:1000), CDK7 (Cell Signaling, #2916, 1:1000), CDK9 (Cell Signaling, #2316, 1:1000), CDK13 (LSBio #LS-B14812, 1:1000), vinculin (Cell Signaling, #13901, 1:1000), RPA32 total (Cell Signaling, #2208, 1:1000), RPA32 pS33 (Cell Signaling, #10148, 1:1000), RPA32 pS4/8 (Bethyl, A300-245A-M, 1:1000), RNAPII total (Cell Signaling, #14958, 1:2000), RNAPII pS2 (Cell Signaling, #13499, 1:5000), RNAPII pS2 (Active Motif, #61699, 1:3000), Histone H3 (Cell Signaling, #9715, 1:1000), and FLAG (Sigma, F1804, 1:1000).

### S9.6 slot blot

Total genomic nucleic acids were purified by lysing cells with standard Proteinase K/SDS/Tris-EDTA lysis buffer followed by standard phenol/chloroform/isoamyl alcohol pH 8.0 extraction and ethanol precipitation. Samples were digested overnight using a restriction enzyme cocktail of BsrgI, EcoRI, HindIII, SspI, and XbaI in Buffer r2.1 (NEB), followed by a 30-minute incubation with RNase A (Thermo Scientific) in 500mM NaCl to remove free RNA. Genomic DNA was extracted as before and precipitated in isopropanol/NaOAc. As a negative control, 8 μg of each sample was treated with RNase H1 (NEB) at 37°C overnight and then purified as before. 500ng of each sample was spotted onto two wells of positively charged Nylon membrane (Roche) using a slot blot apparatus (BioRad) and vacuum suction. As a separate loading control, half of the membrane was denatured in 0.5M NaOH, 1.5M NaCl, then neutralized for 10 minutes in 1M NaCl, 0.5M Tris-HCl pH 7.0. Membranes were UV crosslinked (0.12J/m^2^) and then blocked in 5% milk/TBST. Membranes were incubated overnight with either S9.6 (Kerafast #ENH001) or ssDNA (Millipore #MAB3868) antibodies, then washed and incubated with anti-mouse secondary-HRP antibodies (Cell Signaling, #7076) for 1 hour at RT before exposure to ECL.

### Experimental detection of transcription-replication collisions

The proximity ligation assay (PLA) was carried out as previously described ^75^. Briefly, cells were seeded at 50,000 cells/well in Ibidi u-Slide 8-well slides (#80806) pre-coated with poly-lysine. Cells were allowed to adhere overnight. The next day, DRB (5,6-Dichlorobenzimidazole 1-β-D-ribofuranoside) was added to wells as a negative control and incubated for 4 hours. Then, cells were washed twice with PBS and then treated with CSK extraction buffer (0.2% Triton X-100, 20 mmol/L HEPES-KOH pH 7.9, 100 mmol/L NaCl, 3 mmol/L MgCl_2_, 300 mmol/L sucrose, 1 mmol/L EGTA) on ice for 3 minutes. Cells were fixed on ice with 4% paraformaldehyde, then ice-cold methanol, and then permeabilized in PBS/0.5% Triton X-100 (PBST) and blocked with 3% BSA in PBS-T for 1 hour. Afterwards, cells were incubated with the indicated primary antibodies (PCNA, Santa Cruz, 1:500; p-RNAPII Ser2, Novus, 1:1000) overnight at 4°C. The next day, cells were incubated with anti-mouse minus and anti-rabbit plus PLA probes (Sigma, DUO92002 and DUO92004) at 37°C for 1 hour. The PLA reaction was performed using Duolink In Situ Detection Red (Sigma, DUO92008), according to the manufacturer’s instructions. Cells were stained with DAPI after completing the washing steps. Images were captured with a Nikon 90i confocal microscope, and the PLA signal was quantified using CellProfiler as previously described ^75^. Statistical testing was performed using Prism software (GraphPad 9) and *P*<0.05 was considered statistically significant after correcting for multiple comparisons using either ANOVA or Kruskal-Wallis test as described in the figure caption.

### Drug dose-response assays

Cells were seeded in a 96-well plate at a density of 1,000 cells per well overnight. The next day, drugs including MK-1775, ceralasertib/AZD6738, CCT245737, AZD7762, prexasertib, CHIR-124 (all from MedChemExpress), Debio 0123, ZN-c3, VE-821, PD0166285, and VX-970 (all from Selleckchem) or DMSO was added. Cells were exposed to the drug for 10-15 days, and cell confluency or nuclei count was measured using an IncuCyte S3 (Sartorius).

### Cell cycle analysis

Cells were counted and plated and allowed to adhere overnight. The next day, BrdU was pulsed into the media for approximately 1 hour and then cells were collected, fixed according to protocol, and stained with BrdU-FITC and FxCycle Violet (Thermo Fisher) according to manufacturer’s protocol (BioLegend Phase Flow kit or Thermo Fisher Click-iT EdU kit). Cells were run on an Attune NxT cytometer (Thermo Fisher) in duplicate, and data were analyzed using FlowJo software (BD) to determine G1, S, and G2/M populations.

### Immunofluorescence for RPA32 foci

Staining for RPA32 in S-phase populations was performed in conjunction with the Click-iT EdU Imaging kit (Invitrogen), with some modifications. Cells were plated in a 96-well plate and allowed to rest overnight, then incubated with 20 μM EdU for 30 minutes. Next, cells were washed with PBS and incubated with or without 2mM hydroxyurea for 4 hours. Cells were pre-extracted with 0.5% NP40, fixed in 4% paraformaldehyde, permeabilized in 0.5% Triton X-100, and washed in 3% BSA/PBS. The “Click” reaction proceeded as per the kit protocol, followed by blocking and staining with an RPA32/RPA2 antibody (Abcam) for 1 hour. After subsequent washes, cells were stained with goat anti-mouse IgG Alexa Fluor 647 (Invitrogen), washed, stained with DAPI and imaged. Plates were imaged using the IN Cell Analyzer 6500HS High Content Analysis System (Cytiva). For quantification of EdU intensity and RPA32 foci count, CellProfiler software was used. Our pipeline generated nuclear masks based on DAPI staining, gated cells by positive or negative EdU based on the threshold of a no-EdU control, and then the number of RPA32 foci across each individual nucleus was quantified. Triplicate wells were analyzed with at least 15 fields per well.

## Acknowledgements

We thank the Center for Research Informatics at the University of Chicago for providing the computing infrastructure. We thank Jace Chen for assistance in artwork, Dr. Kari Herrington for microscopy assistance, and the Nikon Imaging Center at UCSF. The work was supported by the National Institutes of Health grant R01CA269977 (L.Y.) and the University of Chicago and UChicago Comprehensive Cancer Center (L.Y.). The work was also supported by Young Investigator Awards from the Prostate Cancer Foundation (to J.C. and H.L.), a Challenge Award from the Prostate Cancer Foundation (to F.Y.F.), the Department of Defense Physician’s Research Award (W81XWH-20-1-0136 to J.C.), as well as funds from the Benioff Initiative for Prostate Cancer Research at UCSF (to J.C., M.K., F.Y.F. and A.A.) and the Martha and Bruce Atwater Breast Cancer Research Program at UCSF (to M.K.).

## Disclosure

A. Ashworth reports personal fees from Tango Therapeutics, Azkarra Therapeutics, Ovibio, Kytarro, Cytomx, Cambridge Science Corporation, Genentech, Gladiator, Circle, Bluestar, Earli, Ambagon, Trial Library, Phoenix Molecular Designs, GSK, Prolynx; grants from SPARC, and AstraZeneca outside the submitted work; in addition, he holds patents on the use of PARP inhibitors held jointly with AstraZeneca from which he has benefited financially (and may do so in the future).

F.Y. Feng reports personal fees from Bluestar Genomics, Astellas, Foundation Medicine, Exact Sciences, Tempus, POINT Biopharma, Janssen, Bayer, Myovant, Roivant, SerImmune, Bristol Meyers Squibb, Novartis, and personal fees from POINT Biopharma outside the submitted work and other support from Artera.

J. Chou reports consulting fees from Exai Bio outside the submitted work.

## Notes

### Funding Statement

The work was supported by the University of Chicago and UChicago Comprehensive Cancer Center (L.Y.). The work was also supported by Young Investigator Awards from the Prostate Cancer Foundation (to J.C. and H.L.), a Challenge Award from the Prostate Cancer Foundation (to F.Y.F.), the Department of Defense Physician's Research Award (W81XWH-20-1-0136 to J.C.), as well as funds from the Benioff Initiative for Prostate Cancer Research at UCSF (to J.C., M.K., F.Y.F. and A.A.) and the Martha and Bruce Atwater Breast Cancer Research Program at UCSF (to M.K.).

### Author Declarations

All data are publicly available prior to this manuscript.

### Summary of Updates

New analyses and new experiments were performed.

