## Supplementary Figures for "Large tandem duplications in cancer result from transcription and DNA replication collisions"

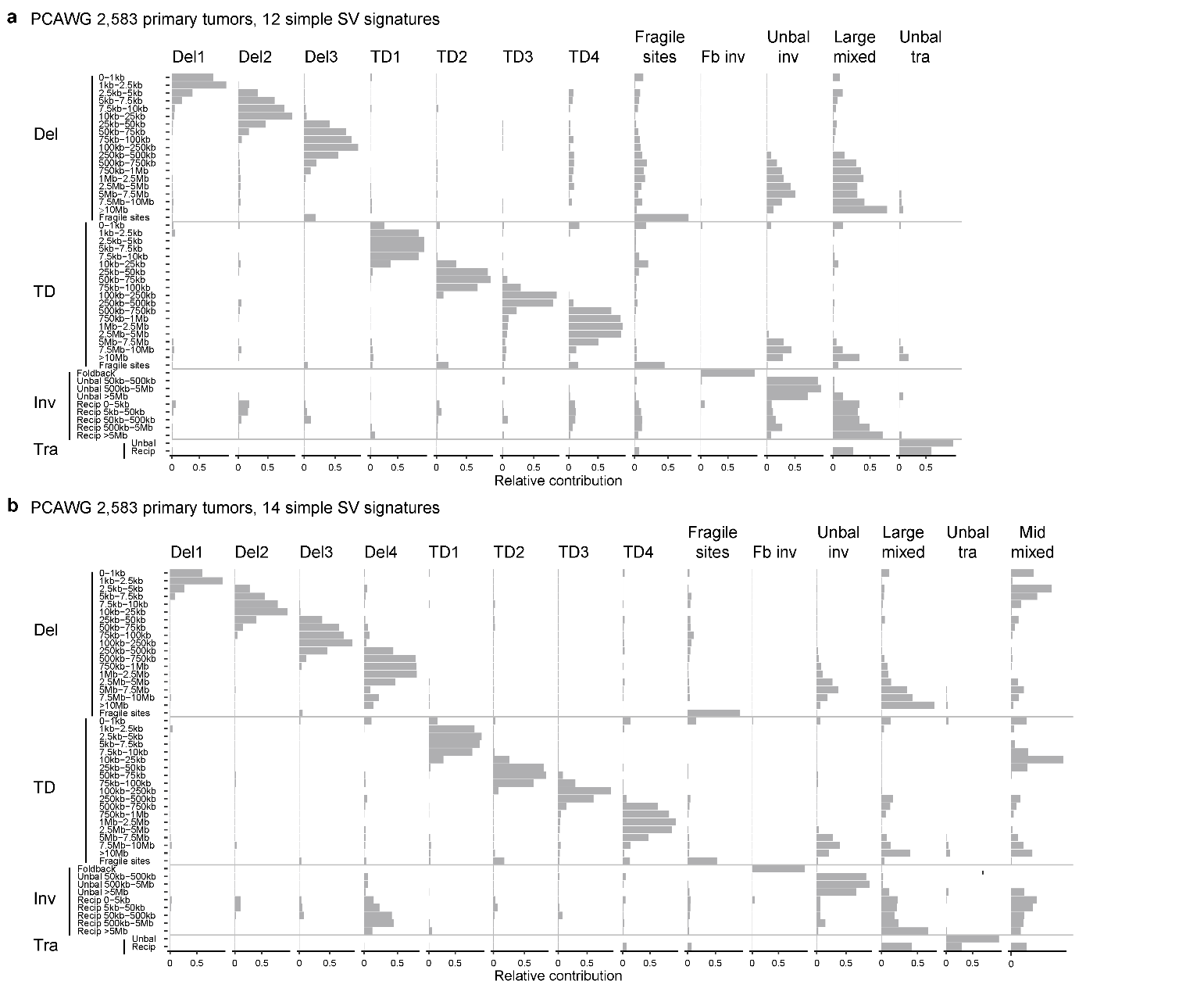

**Supplementary Figure S1. Simple SV signatures in the PCAWG cohort. a** and **b** show 12 and 14 simple SV signatures, respectively. In **a**, there are only three deletion signatures compared to four in **Fig. 1b**. In **b**, there is an additional “Mid mixed” signature compared to the 13 signatures in **Fig. 1b**. The four tandem duplication signatures are unchanged.

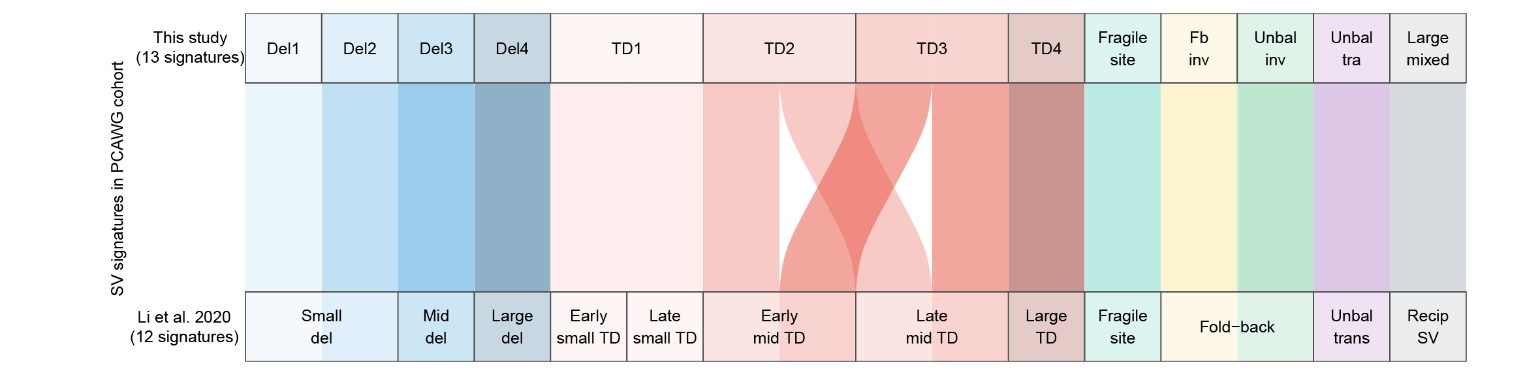

**Supplementary Figure S2. Alluvial plot showing the comparison of simple SV signatures between this study and the PCAWG SV signature study ^2^ on the PCAWG cohort.** Del1 and Del2 of this study correspond to small del in the PCAWG study. TD1 corresponds to early and late small TD signatures. TD2 and TD3 are early and late mid TD signatures. Fb inv and Unbal inv signatures correspond to Fold-back signatures. All other signatures have one-to-one matches between the two studies.

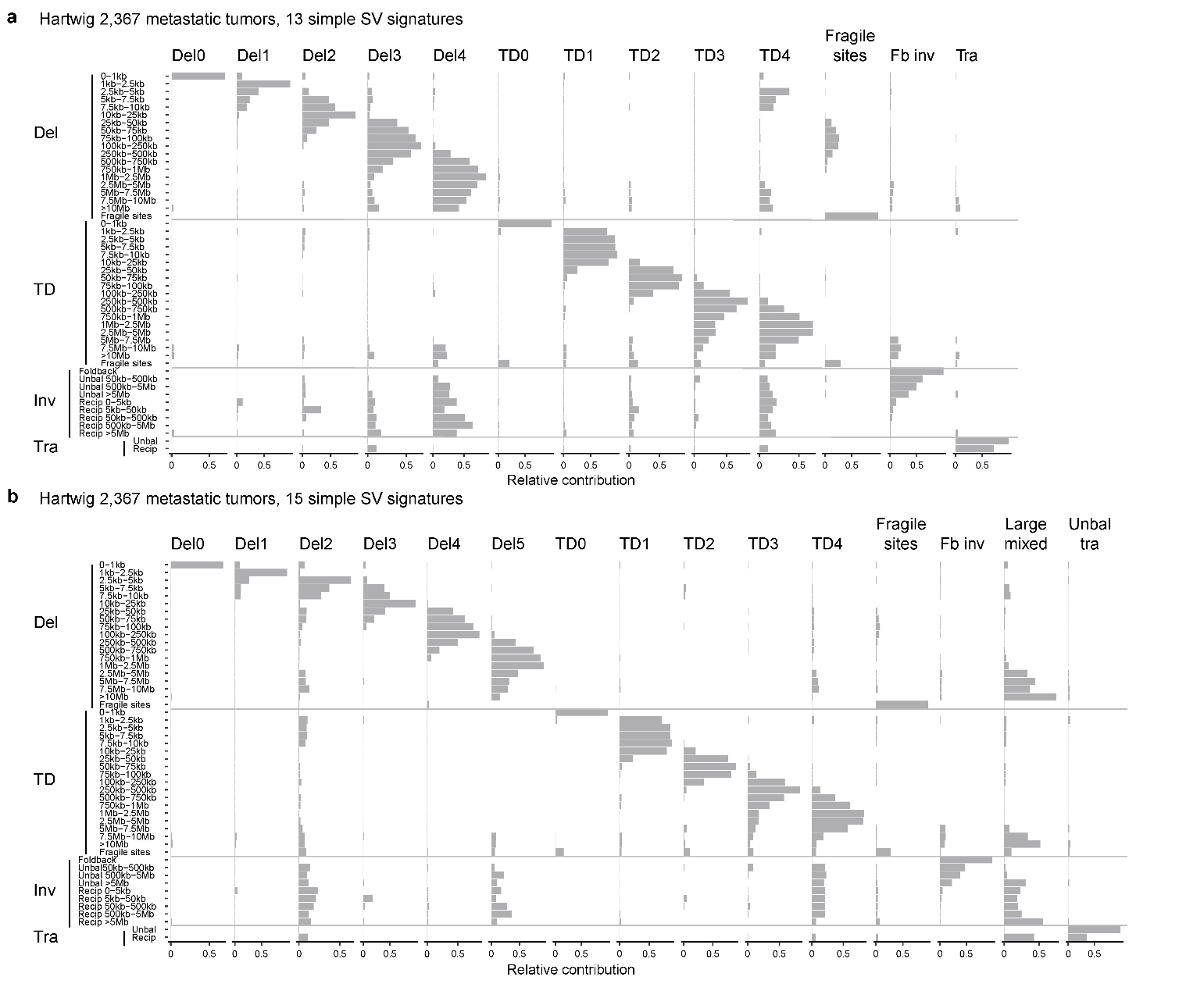

**Supplementary Figure S3. Simple SV signatures in the Hartwig cohort. a** and **b** show 13 and 15 simple SV signatures, respectively. Regardless of the number of signatures selected, there are always five deletion signatures and five tandem duplication signatures, the same as **Fig. 1c**.

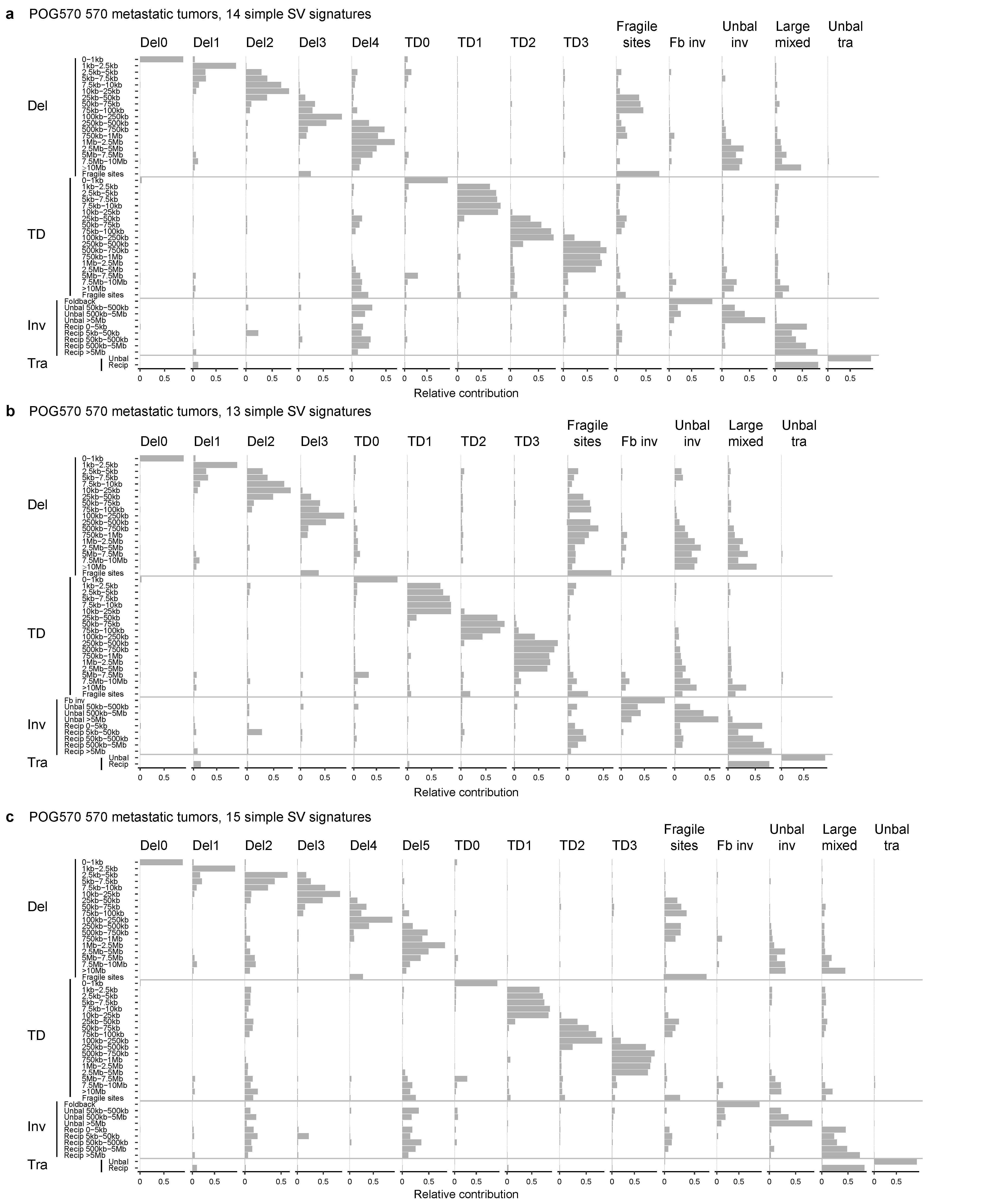

**Supplementary Figure S4. Simple SV signatures in the POG570 cohort. a**, **b**, and **c** show 14, 13 and 15 signatures, respectively. Regardless of the number of signatures selected, there are always four tandem duplication signatures. The only difference is the number of deletion signatures. Compared to the Hartwig cohort, TD3 and TD4 are merged into TD3 in the POG570 cohort.

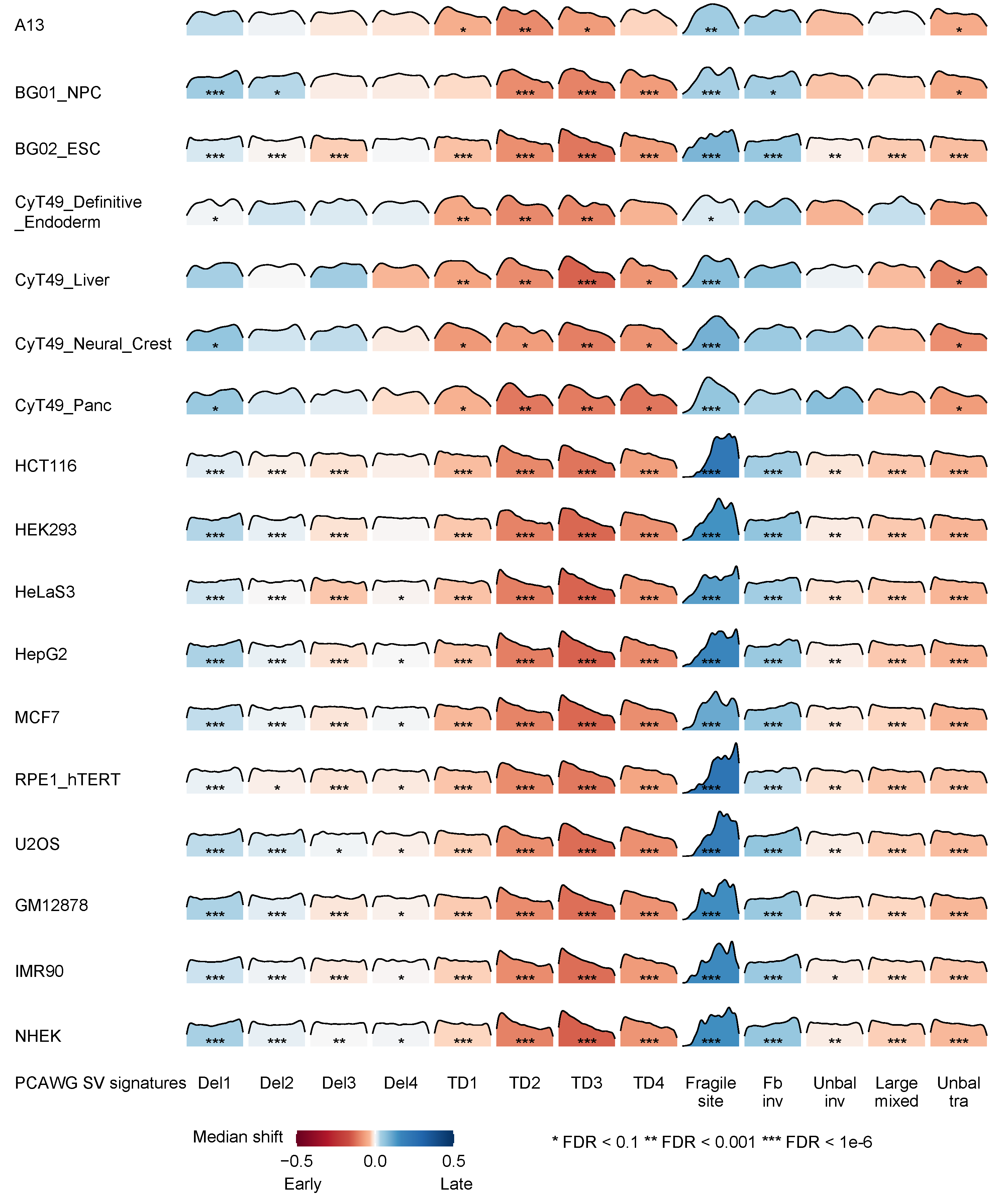

**Supplementary Figure S5. Associations between replication timing and 13 signatures in the PCAWG cohort.** The SVs in 13 signatures from the PCAWG cohort were tested for replication timing in 17 cell lines including the 3 cell lines tested by the PCAWG SV signature study ^2^. Each density curve shows the distribution of replication timing for observed SV breakpoints. The color shows the direction of shift for observed breakpoints in comparison to random breakpoints. Red and blue indicate that observed SV breakpoints are enriched in early and late replicated regions, respectively. A two-sided Kolmogorov-Smirnov test was used to calculate *P* values, and FDR correction was performed.

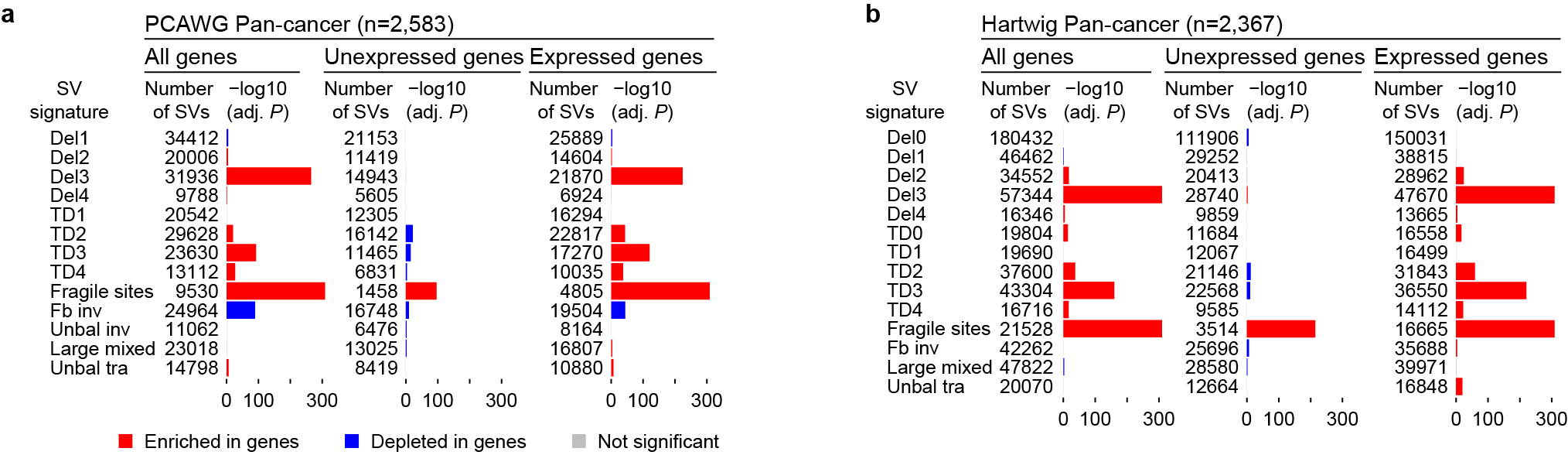

**Supplementary Figure S6. Somatic SVs enriched in genes. a** and **b** show the PCAWG and Hartwig cohorts, respectively. Del3, TD2, TD3, TD4, fragile site and large mixed SVs are enriched in expressed genes in both cohorts. *P* values are calculated by Fisher’s exact test. The x-axis shows Bonferroni-adjusted *P* values.

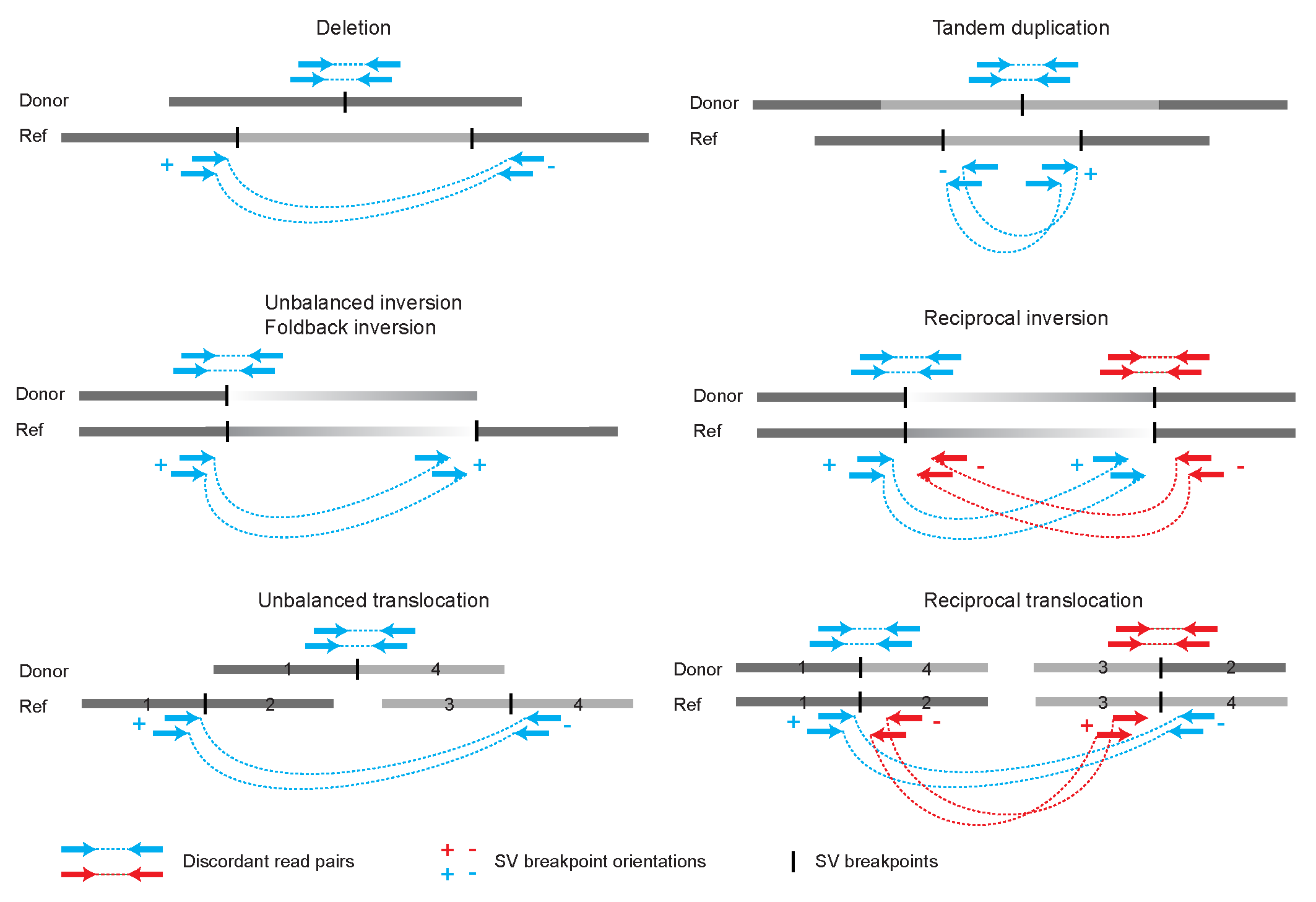

**Supplementary Figure S7. Definition of SV breakpoint orientations.** Reference and donor genomes are shown as grey bars. Blue and red arrows connected by dashed lines are discordant read pairs. The SV breakpoint orientations are defined by the orientations of the discordant reads mapping onto the reference genome.

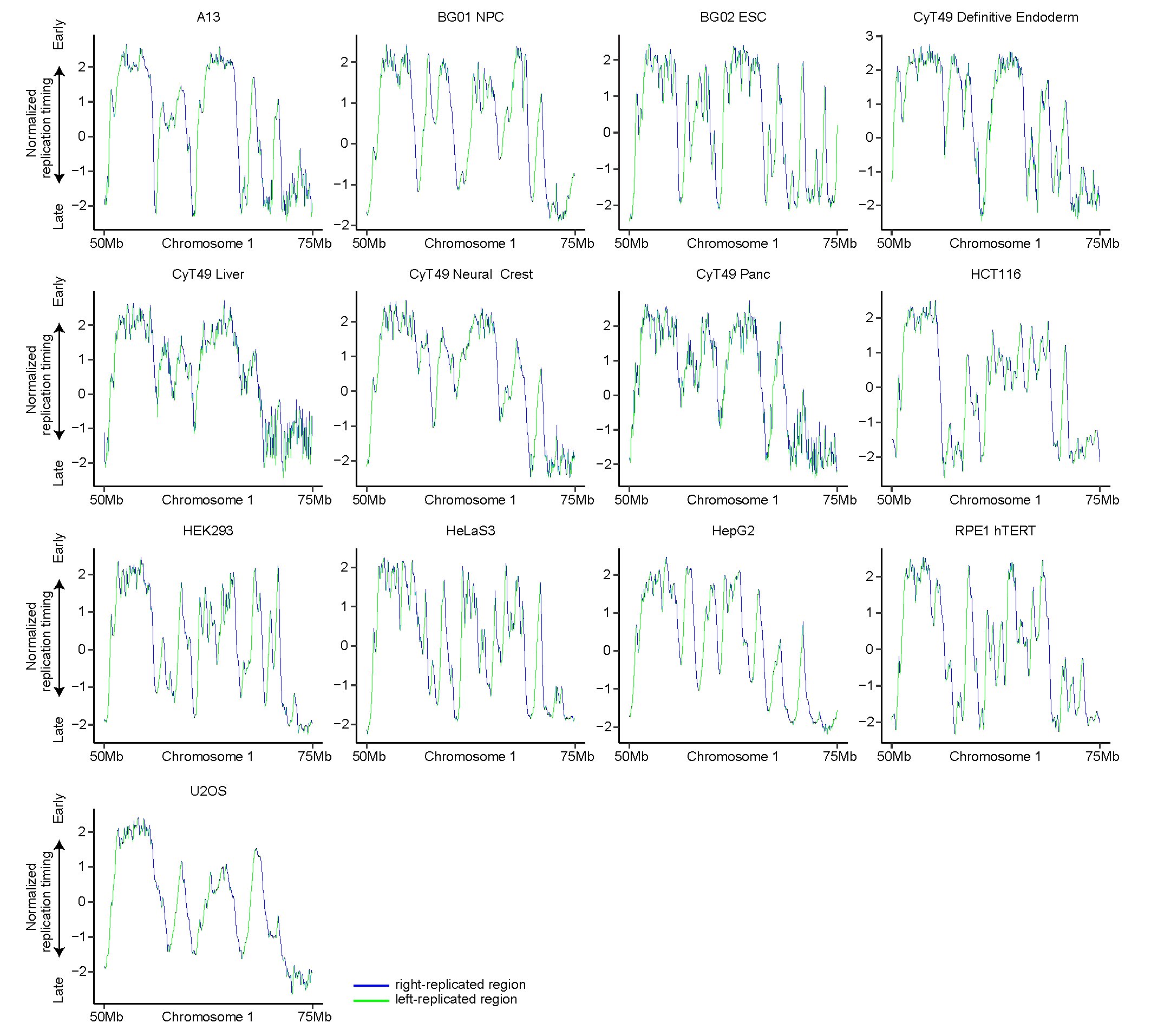

**Supplementary Figure S8. Normalized replication timing in 13 cell lines derived from solid tissues.** The blue and green curves represent right- and left-replicated regions (sharp transition regions).

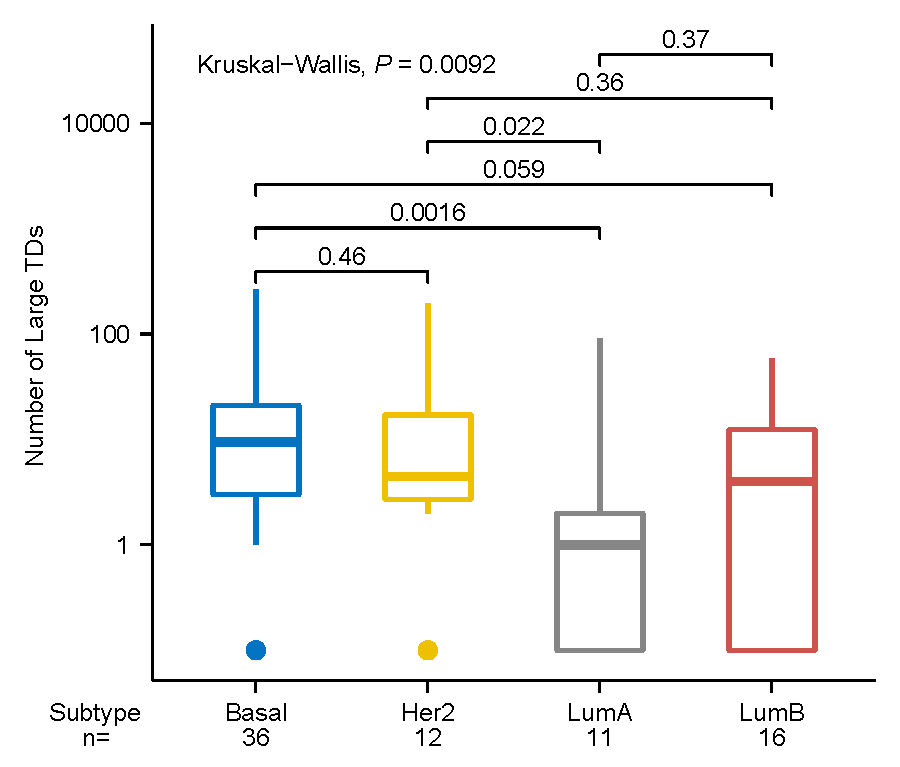

**Supplementary Figure S9. Large TD frequencies in different breast cancer subtypes.** Basal and Her2 types carry more large TDs. *P* values are calculated by Kruskal-Wallis test. The boxplot shows median values (thick colored bars), upper and lower quartiles (boxes), 1.5× interquartile ranges (whiskers), and outliers (colored dots).

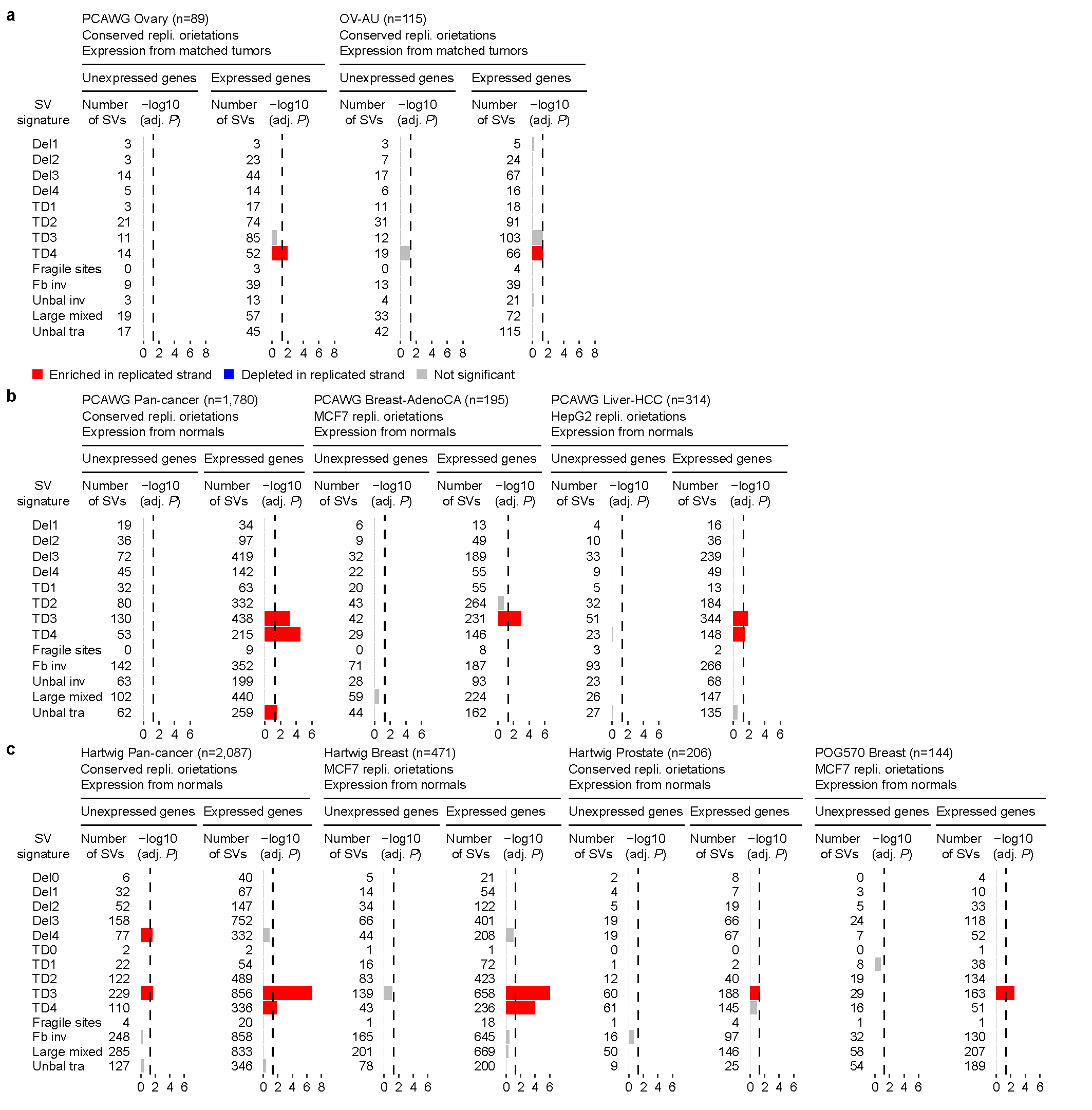

**Supplementary Figure S10. Replicated-strand bias only observed in expressed genes, not in unexpressed genes.** The tested tumor type, sample size, and source of replication orientations are annotated at the top of each plot. Each signature and its number of observed breakpoints are shown on the y-axis. *P* values are calculated by Fisher’s exact test. The x-axis shows Bonferroni-adjusted *P* values.

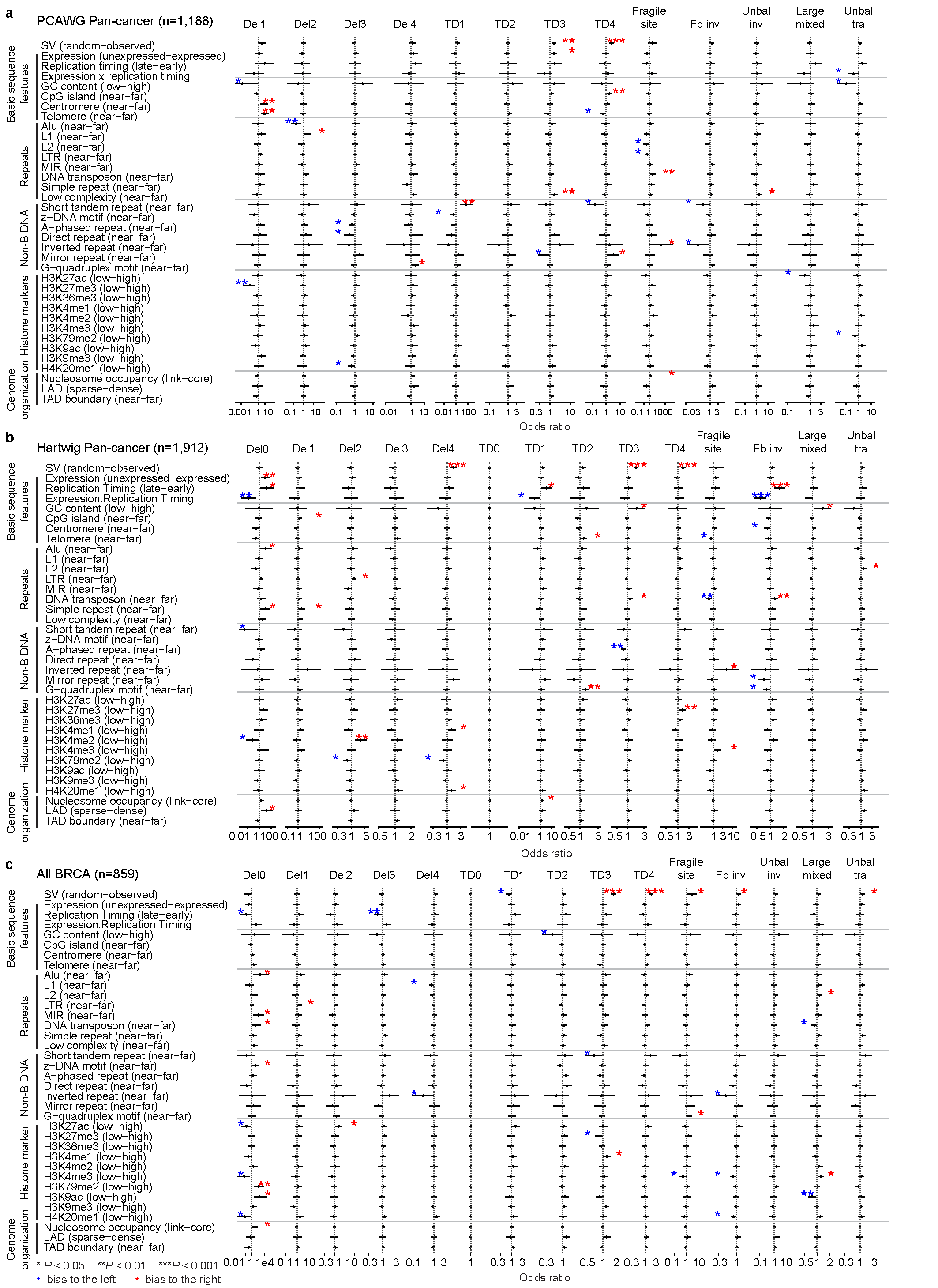

**Supplementary Figure S11. Multi-variate regressions of SV strands and genomic features. a, b** and **c** show the PCAWG cohort, the Hartwig cohort, and the four breast cancer cohorts (PCAWG breast, Hartwig breast, POG570 breast, and BRCA-EU) combined, respectively. One regression is performed for each simple SV signature. Replicated/unreplicated strand is the dependent variable. Genomic features listed on the y-axis are independent variables. Odds ratios are shown on the x-axis with stars indicating significance levels (number of stars) and directions of associations (color of stars). Observed SVs are significantly associated with replicated strands in TD3 and TD4 in the PCAWG cohort, Hartwig cohort, and the combined breast cancer cohort.

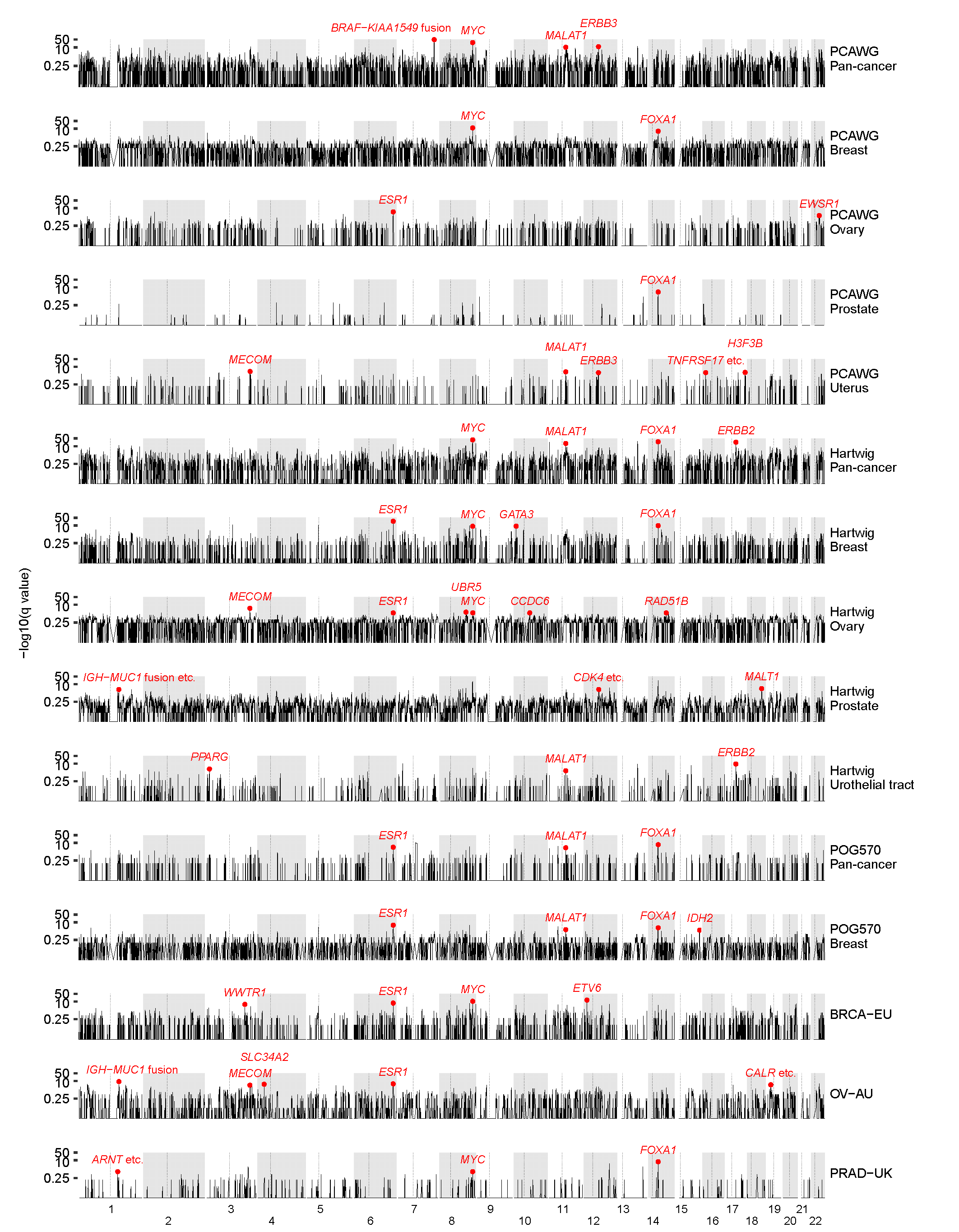

**Supplementary Figure S12. Large TD hotspots in autosomes detected in pan-cancer cohorts, breast, ovarian, prostate, uterus, urothelial-tract cancers, BRCA-EU, OV-AU, and PRAD-UK cohorts.** Known oncogenes are labelled as red dots. Q values are provided by GISTIC2.

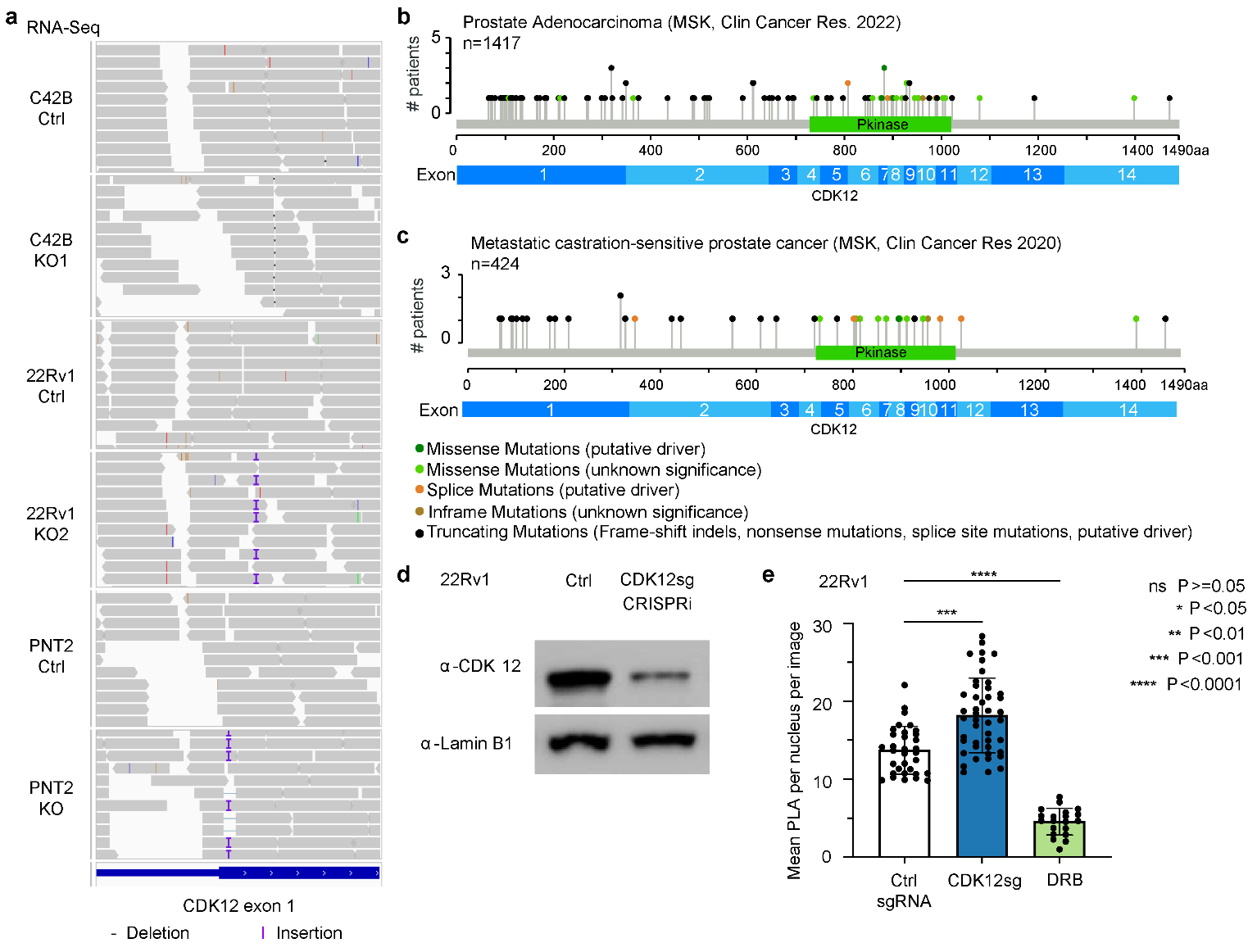

**Supplementary Figure S13. CDK12 KO by CRISPR editing and CDK12 suppression by CRISPR interference (CRISPRi). a**, RNA-Seq reads showing indels in *CDK12* exon 1 in C42B, 22Rv1, and PNT2 Ctrl and KO cells. Grey horizontal bars represent sequencing reads. **b** and **c**, *CDK12* mutations in primary (**b**) and metastatic (**c**) prostate cancers from Memorial Sloan Kettering Cancer Center. **d**, WB showing CDK12 in 22Rv1 Ctrl and CDK12 CRISPRi knockdown (CDK12sg) clone. Lamin B1 is shown as the loading control. **e**, Quantification of the mean PLA signal per nucleus per image in 22Rv1 Ctrl and CDK12sg cells (n=30 fields in one representative experiment). DRB is used as a negative control. Three independent experiments were performed. *P* values are calculated by ANOVA and corrected for multiple tests. Box plots show means (top bounds of boxes) and standard errors of the mean (SEM, error bars). Individual nuclei are shown as black dots.

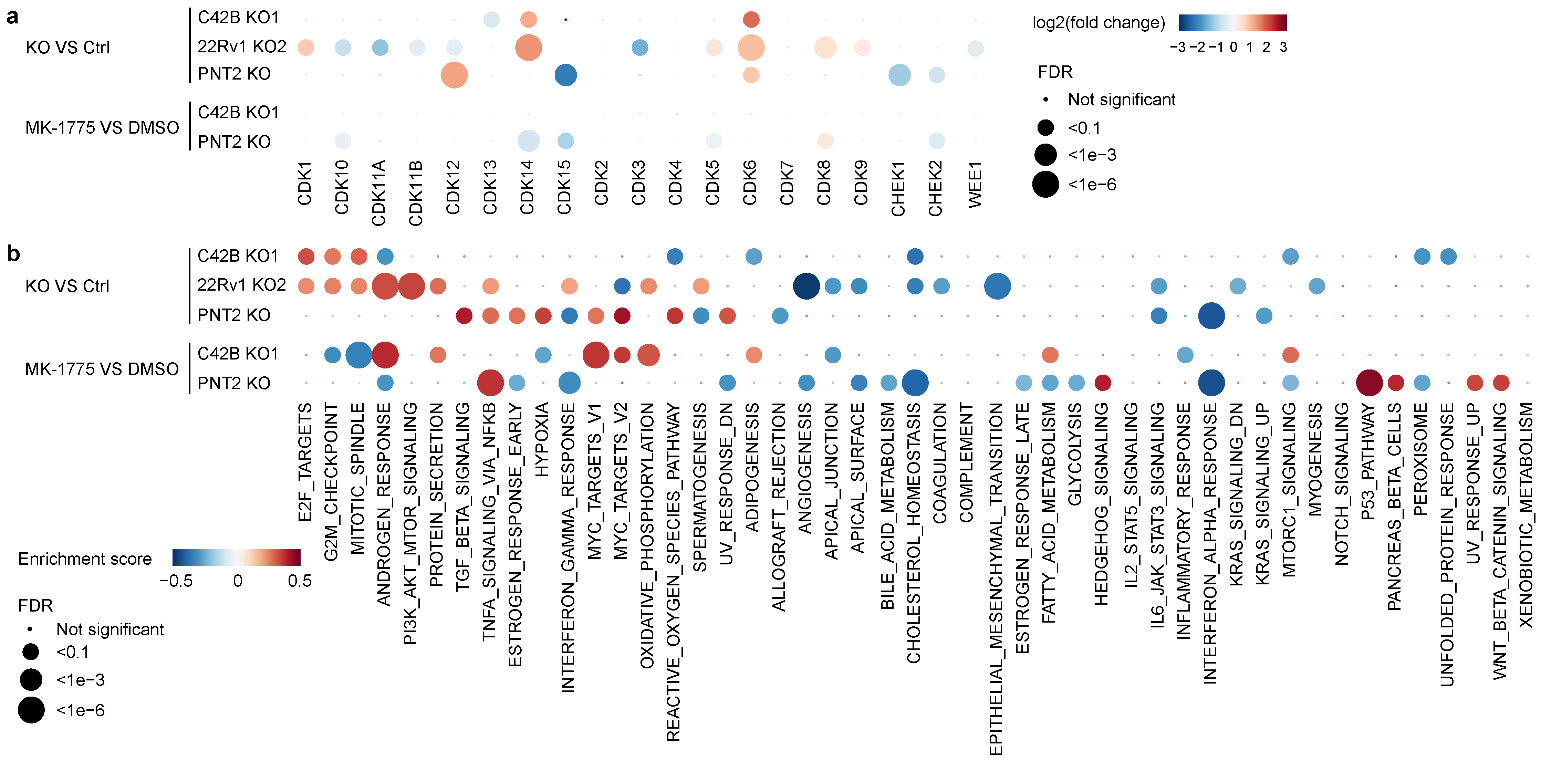

**Supplementary Figure S14. Effects of CDK12 KO in C42B, 22Rv1, and PNT2 cells. a**, Differentially expressed genes. **b**, Gene set enrichment analysis (GSEA). In both **a** and **b**, the comparisons performed are shown on the y axis. Differentially expressed genes and pathways are shown as the x axis. Red and blue dots represent up-regulated and down-regulated genes and pathways, respectively. The sizes of the dots indicate significance levels.

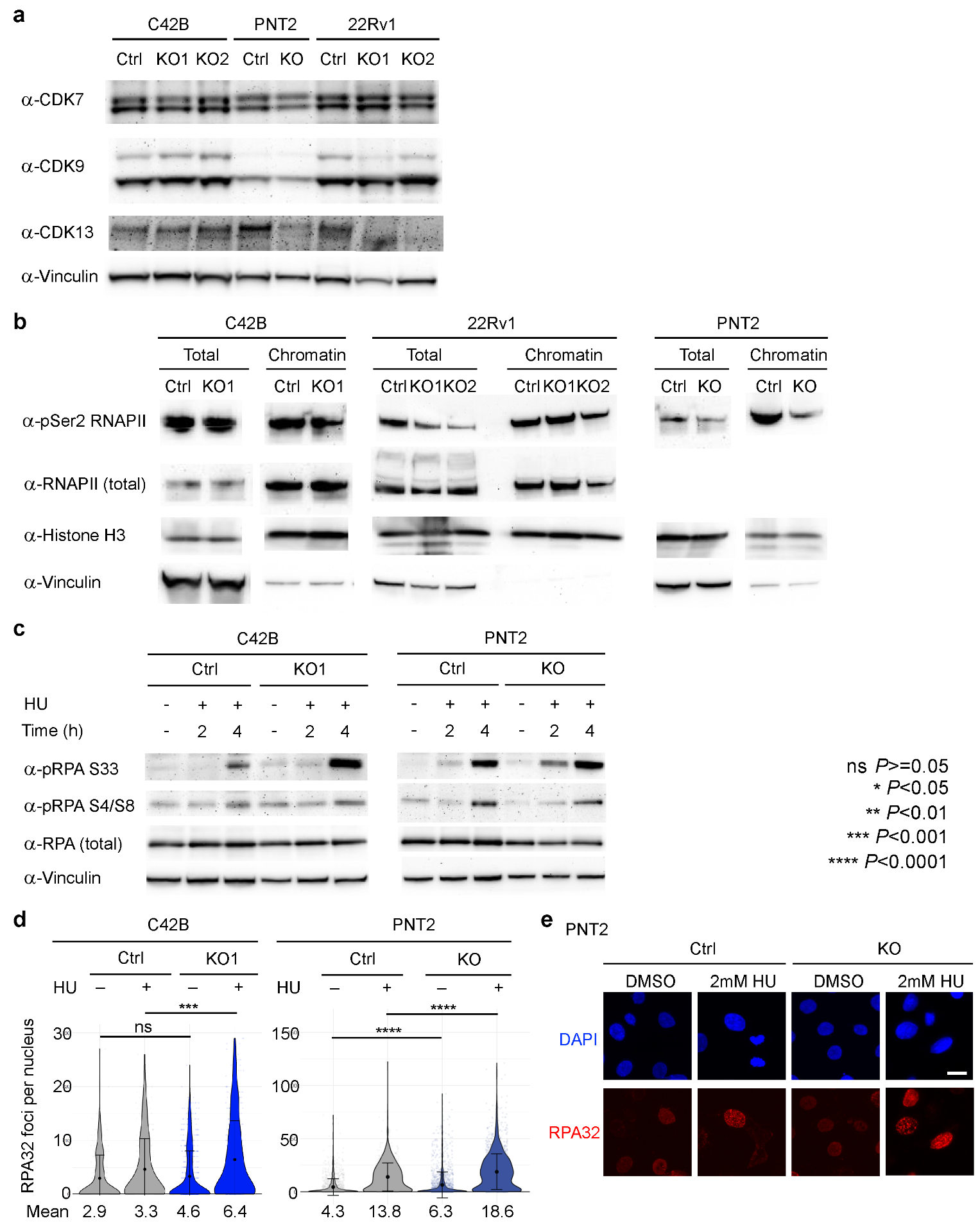

**Supplementary Figure S15. The impact of CDK12 KO on RNAPII and RPA. a**, WB showing CDK7, CDK9, and CDK13 in Ctrl and CDK12 KO cells in C42B, PNT2, and 22Rv1 cell lines. **b**, WB showing RNAPII and Ser2 phosphorylated RNAPII in Ctrl and CDK12 KO cells in C42B, PNT2, and 22Rv1 cell lines. Both whole cell lysate (total) and chromatin fractionation (chromatin) are shown. **c**, WB showing phosphorylated RPA S33, phosphorylated RPA S4/S8, and total RPA in Ctrl and CDK12 KO cells in C42B and PNT2 cell lines treated with or without 2mM hydroxyurea (HU) for the indicated times. **d**, Quantification of the RPA32 foci per nucleus in EduU-positive C42B and PNT2 Ctrl and CDK12 KO cells, with and without HU treatment. (n= at least 790 nuclei [ranging from 790 to 1899 nuclei per condition] of one representative experiment). Two independent experiments were performed. The mean RPA32 foci count per nucleus for each condition is shown at the bottom. **e**, Representative immunofluorescence microscopy images of nuclei (DAPI, blue) and RPA32 foci (red) in Ctrl and CDK12 KO PNT2 cells. Scale bar denotes 20 μm.

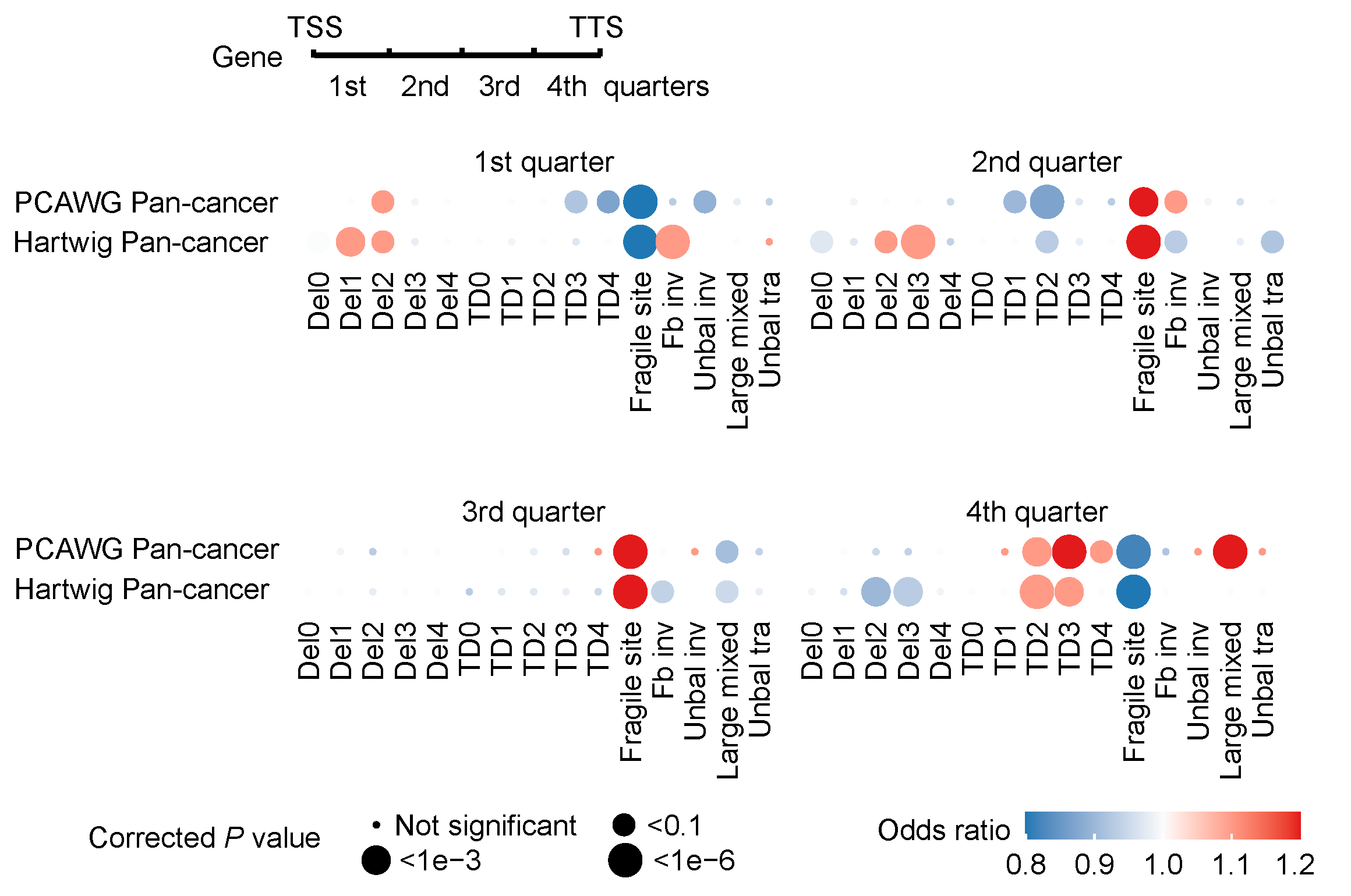

**Supplementary Figure S16. Large TDs are enriched at the end of genes.** Each dot represents the enrichment of the SV signature with respect to the locations of genes. The size of the dot reflects significance levels. Red and blue colors of the dots indicate the enrichment and depletion, respectively. Small deletions and foldback inversions are enriched at the beginning of genes. Fragile site SVs are enriched at the gene bodies. Median and large TDs are enriched at the end of genes. *P* values are calculated by two-sided Fisher’s exact test and Bonferroni correction was performed.

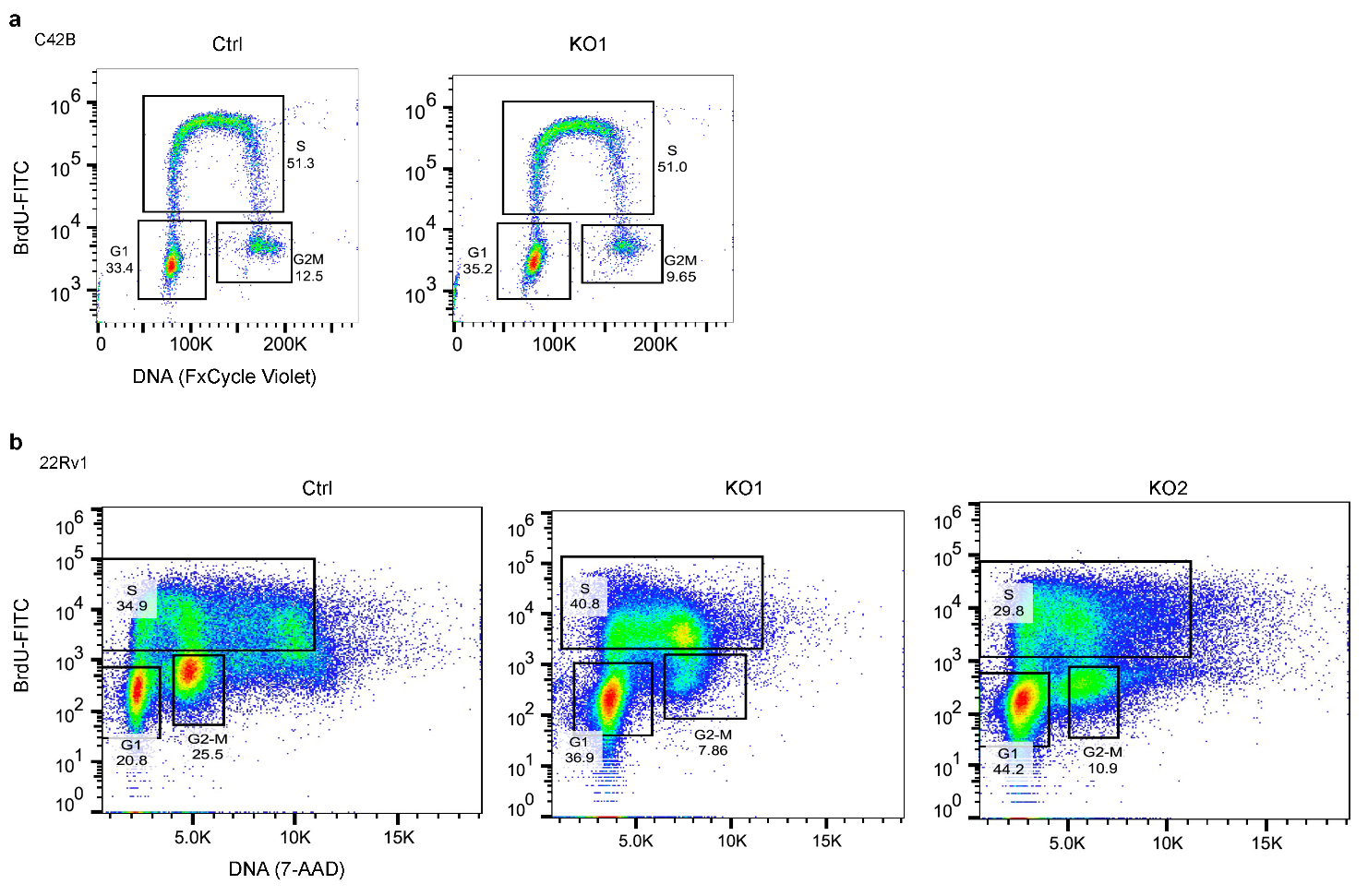

**Supplementary Figure S17. Cell cycle analysis of C42B and 22Rv1 Ctrl and CDK12 KO cells. a**, C42B. **b**, 22Rv1. Cell cycle analysis of C42B (**a**) and 22Rv1 (**b**) Ctrl and CDK12 KO cells. BrdU was pulsed for 1 hour to label cells active in S-phase, processed, and then stained with a BrdU-FITC antibody. The DNA was labeled with either FxCycle violet (**a**) or 7-AAD (**b**). Representative plots from one experiment are shown. A total of 3 independent experiments were performed. Gates for G1, S, and G2-M populations are drawn and the percentage of cells in each gate is shown.

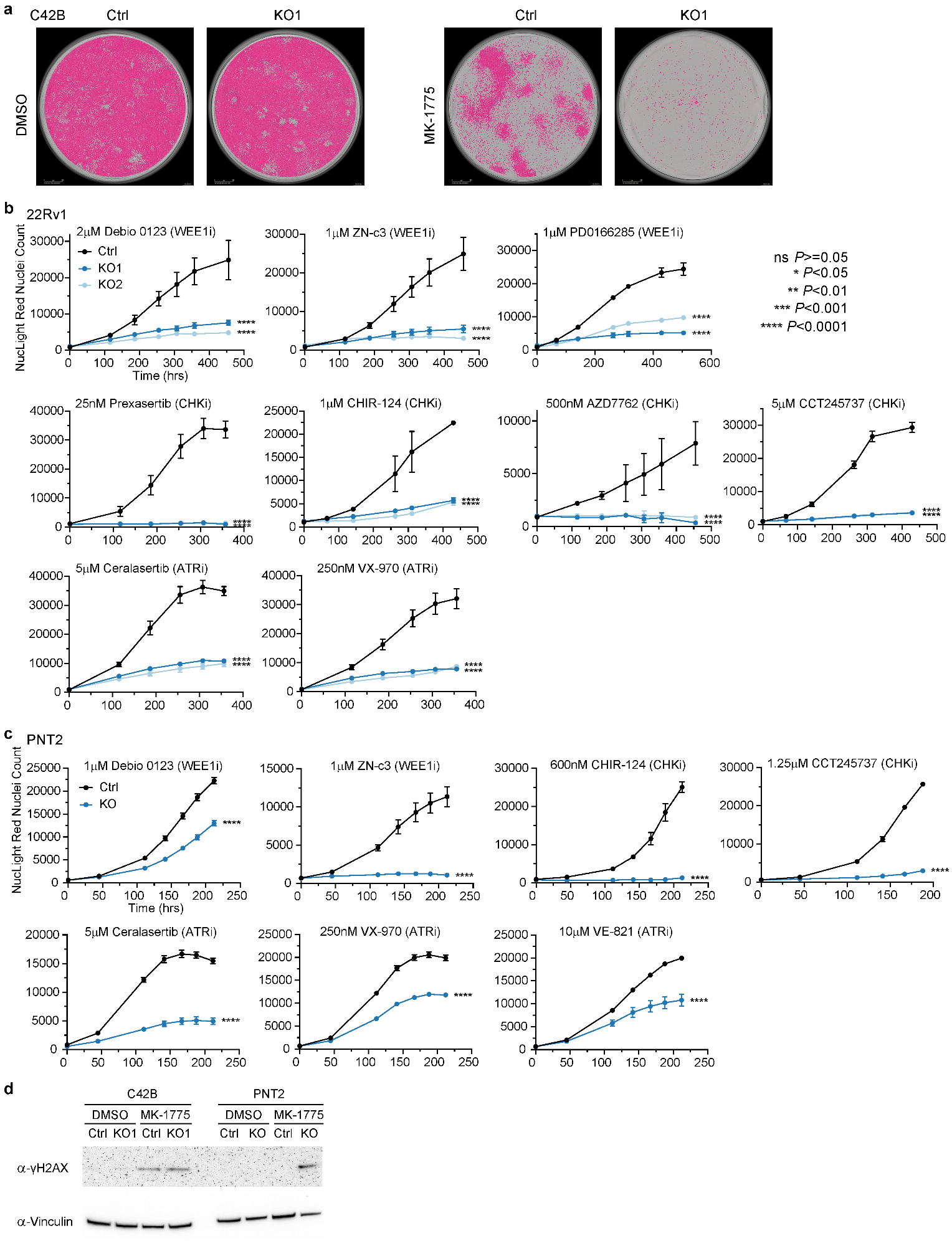

**Supplementary Figure S18. Drug treatment on CDK12 KO cells. a**, Phase contrast images shown with the red nuclei mask (pink) for C42B Ctrl and CDK12 KO cells treated with DMSO or 0.3 mM of MK-1775 on day 12. **b**, Growth curves of 22Rv1 Ctrl and CDK12 KO cells (KO1 and KO2) treated with the indicated drugs (targeting WEE1, CHK1 and ATR) are shown. **C**, Growth curves of PNT2 Ctrl and CDK12 KO cells treated with the indicated drugs (targeting WEE1, CHK1 and ATR) are shown. In both **b** and **c**, error bars represent standard errors of the mean. The *P* value is calculated by Student’s t test for the final day indicated for each experiment. Significant levels are indicated by “*” next to the corresponding CDK12 KO cells compared to Ctrl cells. **d**, WB showing γH2AX in C42B and PNT2 Ctrl and CDK12 KO cells, with and without MK-1775 treatment.
